# Causal Effects of Childhood BMI on Regional Fat Distribution in Adults: A Mendelian Randomisation and Proteomic Mediation Analysis

**DOI:** 10.64898/2026.07.22.26358642

**Authors:** B Hayes, E Hazelwood, G Power, J Gilbody, DJ Pournaras, H Freisling, R Richmond, EE Vincent

**Affiliations:** MRC Integrative Epidemiology Unit, University of Bristol, Bristol, United Kingdom; Translational Health Sciences, Bristol Medical School, University of Bristol, Bristol, United Kingdom; Population Health Sciences, Bristol Medical School, University of Bristol, Bristol, United Kingdom; Early Cancer Institute, University of Cambridge, Cambridge, UK; Francis Crick Institute, London, UK; Institute for Molecular Bioscience, The University of Queensland, Brisbane, Queensland, Australia; Department of Upper GI and Bariatric/Metabolic Surgery, North Bristol NHS Trust, Southmead Hospital, Bristol, United Kingdom; Nutrition and Metabolism Branch, International Agency for Research on Cancer (IARC- WHO), Lyon, France; National Institute for Health and Care Research, Oxford Biomedical Research Centre, John Radcliffe Hospital, Oxford, UK

## Abstract

Childhood obesity is a major global health concern, yet the long-term effects of early-life adiposity on adult fat distribution and underlying biological pathways remain poorly understood. In particular, it is unclear whether childhood BMI differentially influences specific adipose tissue depots and whether circulating proteins mediate these relationships.

We conducted a Mendelian randomisation study using genetic instruments for childhood BMI across 12 timepoints from ages 3 to 18 years. Two-sample MR was applied to five MRI-derived adult fat depots (abdominal subcutaneous (ASAT), gluteofemoral (GFAT), visceral (VAT), liver, and pancreatic fat) in UK Biobank. We further implemented a two-step MR framework to assess whether 2,940 circulating plasma proteins (Olink) mediate observed associations, integrating temporal, statistical, and directional evidence across childhood.

Genetically predicted higher childhood BMI from approximately age 7 years onwards was associated with increased ASAT and GFAT in adulthood, but showed little evidence of association with visceral, liver, or pancreatic fat, with effects persisting into adolescence. Two-step MR identified 140 proteins influenced by childhood BMI during a developmentally sensitive window, of which seven showed directionally consistent evidence of mediation. These proteins included ACAN, CCL7, and CLIC5 for abdominal subcutaneous fat, and CLIC5, BMP10, CLEC10A, KIT, and IGSF3 for gluteofemoral fat, highlighting partially distinct biological pathways across depots.

Childhood BMI exerts depot-specific effects on adult fat distribution, particularly influencing subcutaneous adipose tissue. Circulating proteins provide evidence of potential mediating pathways, supporting the existence of developmentally sensitive biological mechanisms linking early-life adiposity to adult body composition. These findings underscore childhood as a critical period for shaping long-term adipose tissue distribution and highlight potential molecular targets for future intervention strategies.

**Author Summary:** How much fat someone carries as a child can shape their health for decades, but where that fat ends up in the body as an adult may matter just as much as how much there is. Fat stored under the skin behaves very differently, biologically, from fat stored around internal organs or within the liver — and these different patterns are linked to different health risks later in life. In this study, we used genetic data to test whether having a higher body weight in childhood causally influences how fat is distributed across the body in adulthood, rather than simply relying on correlations that could be explained by other factors. We also looked at levels of specific proteins in the blood to see whether they might help explain this link — acting as a biological bridge between childhood body weight and where fat settles later in life. By pinpointing proteins that may carry this influence forward, we hope to shed light on the biological pathways connecting early-life body weight to long-term patterns of fat distribution, which could inform future research into prevention or intervention strategies. This work is part of a broader research programme investigating how body fat distribution relates to health outcomes across the life course.

## INTRODUCTION

Childhood obesity is a critical public health issue, with its prevalence escalating globally over recent decades [1]. Among children and adolescents aged 5-19 years, prevalence of overweight and obesity has risen from ∼4% in 1975 to nearly 20% in 2022, representing a fivefold increase [2]. Between 1990 and 2021, childhood obesity prevalence tripled, with over 93 million children aged 5-14 years living with obesity by 2021 [3]. Excess adiposity during early life is widely recognised as an important determinant of adverse health effects in adulthood, including insulin resistance, increased cardiovascular risk, and cancer [4–6].

With the increasing prevalence of childhood obesity worldwide, understanding the long-term consequences of excess adiposity in early life has become increasingly important. Additionally, the accumulation of adipose tissue in different anatomical sites is thought to play distinct roles in the pathophysiology of numerous health outcomes. Evidence suggests that greater accumulation of visceral and ectopic fat depots is associated with adverse metabolic health, whereas gluteofemoral fat accumulation may confer metabolic benefits relative to fat stored in visceral or ectopic depots[7,8]. The relationship between childhood body mass index (BMI) and regional fat distribution in adulthood, particularly with respect to adipose tissue depots such as abdominal subcutaneous adipose tissue (ASAT), gluteofemoral adipose tissue (GFAT), visceral adipose tissue (VAT), liver fat, and pancreatic fat, remains incompletely understood.

Beyond anthropometric measures, adipose tissue is increasingly recognised as a dynamic endocrine organ that secretes a wide range of bioactive proteins, including adipokines, cytokines, growth factors, and metabolic regulators, such as leptin, adiponectin, interleukin-6 (IL-6), tumour necrosis factor-α (TNF-α), and insulin-like growth factor binding proteins (IGFBPs) [9,10]. These circulating proteins may represent key biological pathways through which excess adiposity in childhood exerts long-lasting effects on tissue-specific fat deposition and metabolic health in adulthood.

Traditional observational studies have provided valuable insights into the associations between adiposity and health outcomes [11–17]. However, these studies have generally examined BMI or regional fat depots as static exposures, rather than investigating whether adiposity at different stages of childhood and adolescence influences the distribution of fat across specific depots in adulthood. While BMI serves as a convenient proxy for overall adiposity, it does not distinguish between fat and lean mass, nor does it capture variation in fat distribution across anatomical depots.

As a result, measures of regional fat distribution provide greater biological and clinical insight beyond BMI alone. This distinction has gained renewed relevance in light of recent publications regarding clinical obesity [18], which proposed redefining obesity based on objective measures of excess adiposity and its functional consequences, rather than BMI thresholds alone. The Commission highlighted that BMI is an imperfect proxy that conflates distinct adiposity phenotypes with meaningfully different health implications, reinforcing the need to characterise fat distribution across specific depots when evaluating metabolic risk.

Additionally, observational analyses are often susceptible to confounding and reverse causality, limiting the ability to infer causality [19,20]. Mendelian randomisation (MR), which exploits the random allocation of genetic variants at conception, and then uses these as proxies for exposures of interest, offers a robust tool to address these limitations by reducing confounding and providing stronger evidence for causal relationships, provided that strong assumptions hold [21,22].

In this study, we use MR to investigate the causal effect of childhood BMI (12 timepoints from 3 to 18 years) on adult fat distribution measures, specifically ASAT, GFAT, VAT, liver fat, and pancreas fat. In addition, we systematically assess the role of circulating proteins in mediating these relationships by analysing 2,940 plasma proteins measured using the OLINK proteomics platform [23]. Specifically, we evaluate whether genetically predicted childhood BMI influences protein expression profiles and whether these proteins, in turn, mediate the effects of childhood adiposity on adult fat deposition, using a two-step MR approach [24].

By leveraging genetic instruments associated with BMI during critical developmental periods, and integrating large-scale proteomic data, this study aims to clarify whether childhood adiposity causally influences the accumulation of fat in potentially harmful depots, and to identify proteomic pathways through which these effects may operate. Understanding these long-term relationships between early-life BMI, tissue-specific fat distribution, and protein pathways could provide important insights into the developmental origins of metabolic disease and inform both preventive and treatment strategies targeting childhood obesity to reduce future disease risk. Preventive strategies might aim to reduce excess adiposity across the childhood population more broadly, while treatment strategies could be more precisely directed at children at highest risk of adverse fat patterning in adulthood — those most likely to benefit from early intervention.

## RESULTS

Participant characteristics across data sources are summarised in **Table 1**. The cBMI exposure data was from ALSPAC, with sample sizes ranging from 736 (age 3 years) to 5,778 (age 10 years) across 12 timepoints [47]. Participants were approximately equally split by sex at younger ages, with a modest female excess at older timepoints (up to 55.4% female at age 18). Median BMI increased across timepoints in both sexes, rising from approximately 16.4–16.5 kg/m² at age 3 to 21.8–22.1 kg/m² at age 18, consistent with expected childhood growth trajectories. Fat depot outcome GWAS were derived from UK Biobank MRI data [31]; participants were predominantly White British, older (mean age ∼64–65 years), and had mean BMI of 26.0 kg/m² in females and 27.1 kg/m² in males. Proteomic data were drawn from the UK Biobank Pharma Proteomics Project (UKB-PPP) [23], with the primary randomised subsample comprising 54,219 participants (mean age 56.7 years, 54.3% female, 94.4% White)

**Table 1.** Participant characteristics across data sources. Values are mean (SD) unless otherwise stated. ALSPAC BMI reported as median (Q1–Q3), sex-stratified. Agrawal et al. age and BMI reported separately by sex. ASAT = abdominal subcutaneous adipose tissue; GFAT = gluteofemoral adipose tissue; VAT = visceral adipose tissue; UKB-PPP = UK Biobank Pharma Proteomics Project

|  | **ALSPAC**<br>**(cBMI**<br>**exposures)** | **Agrawal et al.**<br>**(ASAT, GFAT,**<br>**VAT)** | **Liu et al.**<br>**Liver fat**<br>**(GWAS cohort)** | **Liu et al.**<br>**Pancreas fat**<br>**(GWAS cohort)** | **UKB-PPP**<br>**(Proteins)** |
| --- | --- | --- | --- | --- | --- |
| **N** | 736–5,778<br>(varies by<br>timepoint) | 39,076 | 32,858 | 25,617 | 54,219 |
| **Age**<br>**(years)** | 3–18<br>(median per<br>timepoint) | F: 63.9 (7.5)<br>M: 65.2 (7.7) | 63.9 (7.52) | 64.2 (7.48) | 56.7 (8.1) |
| **Sex**<br>**(% female)** | 48.5–55.4%<br>across<br>timepoints | 51.3% | 51.5% | 51.2% | 54.3% |
| **BMI**<br>**(kg/m<sup>2</sup>)** | Median ~16.4–<br>22.1<br>across<br>timepoints | F: 26.0 (4.6)<br>M: 27.1 (3.8) | 26.5 (4.36) | 26.5 (4.31) | 27.4 (4.8) |
| **Ancestry** | European | Predominantly<br>White British | 100%<br>White British | 100%<br>White British | 94.4%<br>White |

### Primary MR: Childhood BMI effect on Adult Fat Distribution

Genetically predicted higher childhood BMI (cBMI) from early childhood through adolescence (3-18 years) showed evidence of age-dependent positive associations with adult abdominal and gluteofemoral subcutaneous adipose tissue (ASAT and GFAT), but not with visceral (VAT) or ectopic (liver and pancreas) fat depots (**Figure 2**). Across outcomes, effect estimates in early childhood were generally modest, with stronger and more consistent associations emerging from mid-childhood at age 7 years onwards. For all primary MR results, effect estimates represent the change in

MRI-derived fat depot volume (in standard deviations) per 1 kg/m² increase in genetically predicted childhood BMI.

For adult ASAT, consistent evidence of an association first appeared from age 7 years (beta= 0.13, 95% = 0.05, 0.21) onwards, with effect sizes increasing through late childhood and into early adolescence. The strongest associations in both magnitude and strength of evidence were observed between ages 8 and 14 years (beta at age 8= 0.52, 95% CI= 0.29,0.75; beta at age 14= 0.39, 95% CI= 0.24, 0.54;

**Table 2a, 2b and Supplementary Table 3**). Associations remained positive through late adolescence (ages 15 to 18 years), although effect sizes were modestly attenuated compared to earlier timepoints.

**Table 2a.** Primary MR analysis: IVW results for childhood BMI effect on adult fat deposition. Two-sample MR results to demonstrate effect of childhood BMI (ages 3-18 years) on adult fat deposition (liver and pancreatic fat).

| BMI exposure<br>at age (years) | nsnp | Fstat | Liver fat |  | Pancreas fat |  |
| --- | --- | --- | --- | --- | --- | --- |
|  |  |  | beta(loci, upci) | pval | beta(loci, upci) | pval |
| 3 | 4 | 5.55 | 0.04(-0.02, 0.10) | 2.05E-01 | 0.05(-0.02, 0.11) | 1.80E-01 |
| 5 | 6 | 4.91 | 0.02(-0.09, 0.14) | 6.91E-01 | 0.03(-0.07, 0.13) | 5.58E-01 |
| 6 | 5 | 5.34 | 0.13(-0.01, 0.28) | 7.54E-02 | 0.03(-0.06, 0.12) | 5.62E-01 |
| 7 | 6 | 7.1 | 0.10(-0.05, 0.25) | 1.94E-01 | 0.07(-0.04, 0.18) | 2.18E-01 |
| 8 | 6 | 8.19 | 0.08(-0.08, 0.24) | 3.34E-01 | 0.11(-0.07, 0.29) | 2.18E-01 |
| 10 | 11 | 10.64 | 0.04(-0.08, 0.16) | 5.06E-01 | 0.08(-0.02, 0.19) | 1.14E-01 |
| 12 | 10 | 9.62 | 0.04(-0.09, 0.17) | 5.55E-01 | 0.09(-0.02, 0.20) | 9.85E-02 |
| 14 | 8 | 10.78 | 0.04(-0.10, 0.18) | 5.86E-01 | 0.09(-0.03, 0.20) | 1.36E-01 |
| 15 | 7 | 9.93 | -0.02(-0.18, 0.13) | 7.60E-01 | 0.05(-0.09, 0.19) | 4.76E-01 |
| 16 | 7 | 9.63 | 0.05(-0.04, 0.15) | 2.99E-01 | 0.03(-0.11, 0.17) | 6.80E-01 |
| 17 | 7 | 8.06 | 0.03(-0.15, 0.20) | 7.40E-01 | 0.10(-0.03, 0.22) | 1.17E-01 |
| 18 | 9 | 7.89 | -0.02(-0.14, 0.10) | 7.25E-01 | 0.02(-0.08, 0.13) | 6.54E-01 |

**Table 2b.** Primary MR analysis: IVW results for childhood BMI effect on adult fat deposition. Two-sample MR results to demonstrate effect of childhood BMI (ages 3-18 years) on adult fat deposition (ASAT, GFAT, VAT)., and liver and pancreatic fat).

| cBMI<br>exposure at<br>age (years) | nsnp | Fstat | ASAT |  | GFAT |  | VAT |  |
| --- | --- | --- | --- | --- | --- | --- | --- | --- |
|  |  |  | beta(loci, upci) | pval | beta(loci, upci) | pval | beta(loci, upci) | pval |
| 3 | 5 | 5.54 | 0.12(-0.02, 0.27) | 9.49E-02 | 0.11(-0.01, 0.22) | 6.79E-02 | 0.05(-0.03, 0.13) | 2.08E-01 |
| 5 | 6 | 4.91 | 0.19(0.00, 0.38) | 4.57E-02 | 0.16(0.01, 0.31) | 3.89E-02 | 0.02(-0.10, 0.14) | 7.12E-01 |
| 6 | 5 | 5.34 | 0.06(-0.09, 0.22) | 4.26E-01 | 0.01(-0.15, 0.17) | 9.25E-01 | 0.10(-0.04, 0.24) | 1.78E-01 |
| 7 | 6 | 7.1 | 0.13(0.05, 0.21) | 9.56E-04 | 0.07(0.01, 0.14) | 2.47E-02 | 0.08(-0.05, 0.21) | 2.08E-01 |
| 8 | 6 | 8.19 | 0.52(0.29, 0.75) | 7.87E-06 | 0.42(0.27, 0.56) | 1.32E-08 | 0.16(-0.04, 0.37) | 1.24E-01 |
| 10 | 11 | 10.64 | 0.37(0.22, 0.52) | 1.92E-06 | 0.29(0.17, 0.40) | 1.36E-06 | 0.09(-0.06, 0.23) | 2.40E-01 |
| 12 | 10 | 9.62 | 0.38(0.22, 0.54) | 1.94E-06 | 0.28(0.14, 0.42) | 7.68E-05 | 0.11(-0.03, 0.25) | 1.24E-01 |
| 14 | 8 | 10.78 | 0.39(0.24, 0.54) | 4.29E-07 | 0.30(0.18, 0.42) | 9.22E-07 | 0.11(-0.04, 0.26) | 1.53E-01 |
| 15 | 8 | 9.2 | 0.26(0.04, 0.49) | 1.95E-02 | 0.21(0.04, 0.37) | 1.39E-02 | 0.04(-0.12, 0.20) | 6.23E-01 |
| 16 | 7 | 9.63 | 0.32(0.15, 0.5) | 3.11E-04 | 0.23(0.08, 0.38) | 2.69E-03 | 0.06(-0.09, 0.22) | 4.19E-01 |
| 17 | 7 | 8.06 | 0.34(0.14, 0.55) | 1.11E-03 | 0.24(0.09, 0.39) | 1.75E-03 | 0.09(-0.09, 0.27) | 3.45E-01 |
| 18 | 9 | 7.89 | 0.23(0.06, 0.41) | 8.22E-03 | 0.18(0.05, 0.31) | 5.61E-03 | 0.02(-0.11, 0.16) | 7.26E-01 |

A similar pattern was observed for adult GFAT (**Table 2a, 2b and Supplementary Table 4**). Evidence of an association was weak or absent at younger ages, but emerged by age 7 years (beta =0.08, 95% CI= 0.01, 0.14). This association was strengthened from age 8 years onwards (beta at age 8= 0.42, 95% CI= 0.27, 0.56), with consistent positive associations observed across later childhood and adolescence (ages 10 to 18 years). Across comparable ages, effect sizes for GFAT were generally smaller than those observed for ASAT.

In contrast, there was little evidence that cBMI was associated with adult VAT, liver fat, or pancreatic fat at any of the timepoints (**Supplementary Tables 5-7**).

Sensitivity analyses broadly support the main IVW findings for both ASAT and GFAT (**Supplementary Tables 3 and 4**). However, F-statistics were low to modest (median= 8.13 (range: 4.91–10.78)) across all timepoints, indicating weak instruments. Between ages 3 to 6 years, associations with both outcomes were generally weak or null, and not consistently supported across alternative MR methods (MR Egger, weighted median, simple mode, and weighted mode). MR Egger estimates at these ages were imprecisely estimated, reflecting poor instrument strength. These estimates remained largely unchanged by SIMEX correction.

From age 7 to 14 years, IVW estimates were consistently positive for both outcomes. During this period, weighted median and mode-based estimates were largely concordant with IVW results. MR Egger estimates were somewhat attenuated but directionally consistent, and SIMEX-corrected MR Egger estimates remained comparable with raw MR Egger estimates. At ages 15 to 18 years, positive associations persisted but were less consistent across sensitivity methods, particularly for GFAT. However, both raw and SIMEX-corrected MR Egger estimates remained broadly similar in direction and magnitude. Overall, the concordance between IVW, pleiotropy-robust methods, and SIMEX-corrected Egger across all timepoints supports the robustness of observed associations.

Sensitivity analyses using Radial MR, MR-LASSO, and MR-PRESSO yielded estimates that were largely consistent with the primary IVW results for both ASAT and GFAT. Radial MR showed similar effect sizes before and after outlier removal, and MR-LASSO estimates were generally concordant with IVW, suggesting limited influence of invalid instruments. Although MR-PRESSO identified outliers at several ages, outlier-corrected estimates remained directionally consistent, providing overall reassurance that the findings are robust to pleiotropy, with stronger and more stable evidence observed for ASAT than GFAT.

#### Supplementary analyses

In sex-stratified IVW analyses, associations between cBMI and adult fat depots were broadly consistent in direction with the primary (combined-sex) results for both ASAT (beta at age 8 in females= 0.43, 95% CI= 0.22, 0.64; beta at age 8 in males= 0.61, 95% CI= 0.32, 0.90) and GFAT (beta at age 8 in females= 0.39, 95% CI= 0.22, 0.51; beta at age 8 in males= 0.45, 95% CI= 0.26, 0.63) (**Supplementary Table 8**). Effect estimates were generally stronger and more precisely estimated in females, particularly from mid-childhood onwards, whereas corresponding estimates in males were typically attenuated. This pattern mirrors the greater adiposity accrual and storage in subcutaneous depots observed in females.

Supplementary analyses using MoBa discovery GWAS instruments (for ages 3 to 8 years), which demonstrated substantially stronger instrument strength, with higher F-statistics (∼33–38) compared to those in our main analysis (∼5–11), yielded results that were broadly consistent with the primary IVW findings (**Supplementary Table 9**). For ASAT and GFAT, IVW estimates from MoBa cBMI exposures showed positive associations emerging from mid-childhood, with evidence of stronger effects from age 7 years (ASAT beta= 0.47, 95% CI= 0.30, 0.64; GFAT beta= 0.35, 95% CI= 0.21, 0.50) compared with earlier timepoints (ASAT beta at cBMI age 3= 0.24, 95% CI= - 0.00, 0.49; GFAT beta at cBMI age 3= 0.15, 95% CI= −0.06, 0.37). This temporal pattern mirrors the shift observed in the main analyses, supporting the robustness of the timing of effects and suggesting that findings are unlikely to be driven by weak instrument bias.

In contrast, IVW estimates for VAT, liver fat, and pancreatic fat were generally smaller and imprecise compared to those for ASAT and GFAT, with 95% confidence intervals overlapping the null across timepoints, consistent with the weaker or absent associations observed in the primary analyses.

### Two-step MR: Childhood BMI effect on protein levels

In this step, we examined 2,940 proteins to identify potential mediators of the effect of childhood BMI (cBMI) on adult fat depots. In Step 1, changes in cBMI effects were noted around 7 years of age, therefore three timepoints either side of 7 years were used to define the relevant window. With 7 exposure timepoints and 2,940 outcomes, this resulted in over 20,000 MR analyses (**Supplementary Table 10**). To systematically narrow down potential candidates, we applied a series of filtering criteria.

First, we required evidence of an effect of cBMI on the protein level at any timepoint. This criterion excluded 1,995 proteins with no detectable cBMI influence, with 945 protein candidates remaining. Next, we assessed whether the observed effects aligned with the temporal window identified in Step 1, (three timepoints on either side of 7 years of age). This filter excluded an additional 805 proteins showing sporadic effects at non-consecutive timepoints, or a consistent effect across all timepoints.

The remaining 140 proteins were classified as either “gain of effect” or “loss of effect” candidates based on the pattern of causal effects of cBMI on protein levels. Hierarchical clustering of causal effect estimates for these 140 candidates revealed two broad clusters of proteins, distinguished by the direction of the effect of childhood BMI on circulating protein levels, with the majority showing a positive relationship, and an uptick in the magnitude of effects between the ages of 7 to 8 years (**Figure 3; Supplementary Table 11**). A loss of effect was defined as a protein for which cBMI showed evidence of a causal effect on circulating levels observed up to years 5, 6, or 7, with no observed effect at later timepoints. Conversely, a gain of effect was defined as a protein showing no causal effect of cBMI on circulating levels up to years 5, 6, or 7, with evidence of a causal effect emerging and persisting in all subsequent timepoints. Applying these definitions, we identified 136 gain of effect and 4 loss of effect candidate mediators from the original set of 2,940 proteins.

### Two-step MR: protein effect on ASAT and GFAT

In the second step of the two-step MR framework, the 140 candidate proteins identified as temporally consistent potential mediators of cBMI effect on fat depots, were evaluated for causal effects on adult adiposity depots, specifically ASAT and GFAT. Volcano plots illustrating the causal effects of the 140 candidate proteins on ASAT (left) and GFAT (right) revealed a largely consistent pattern across both depots. A greater number of proteins reached the significance threshold (p < 0.05, dotted line) in the GFAT analysis, with effects predominantly in the negative direction (**Figure 4; Supplementary Table 12**). Effect estimates are reported as standard deviation changes in fat depot volume per genetically predicted standard deviation increase in protein levels.

Evidence consistent with a causal effect on ASAT was observed for 11 candidate proteins (**Table 3**). Among these, an inverse association with ASAT was found for genetically predicted levels of ACAN (beta= −0.15, 95% CI= −0.27, −0.04), PRTN3 (beta= −0.08, 95% CI= −0.12, −0.03), CD300C (beta= −0.06, 95% CI= −0.09, −0.02), COL6A3 (beta= −0.13, 95% CI= −0.25, −0.02), EBI3 (beta= −0.38, 95% CI= −0.58, − 0.17), and SEMA3F (beta= −0.17, 95% CI= −0.27, −0.07), suggesting lower abdominal subcutaneous fat with higher circulating protein concentrations.

**Table 3.**
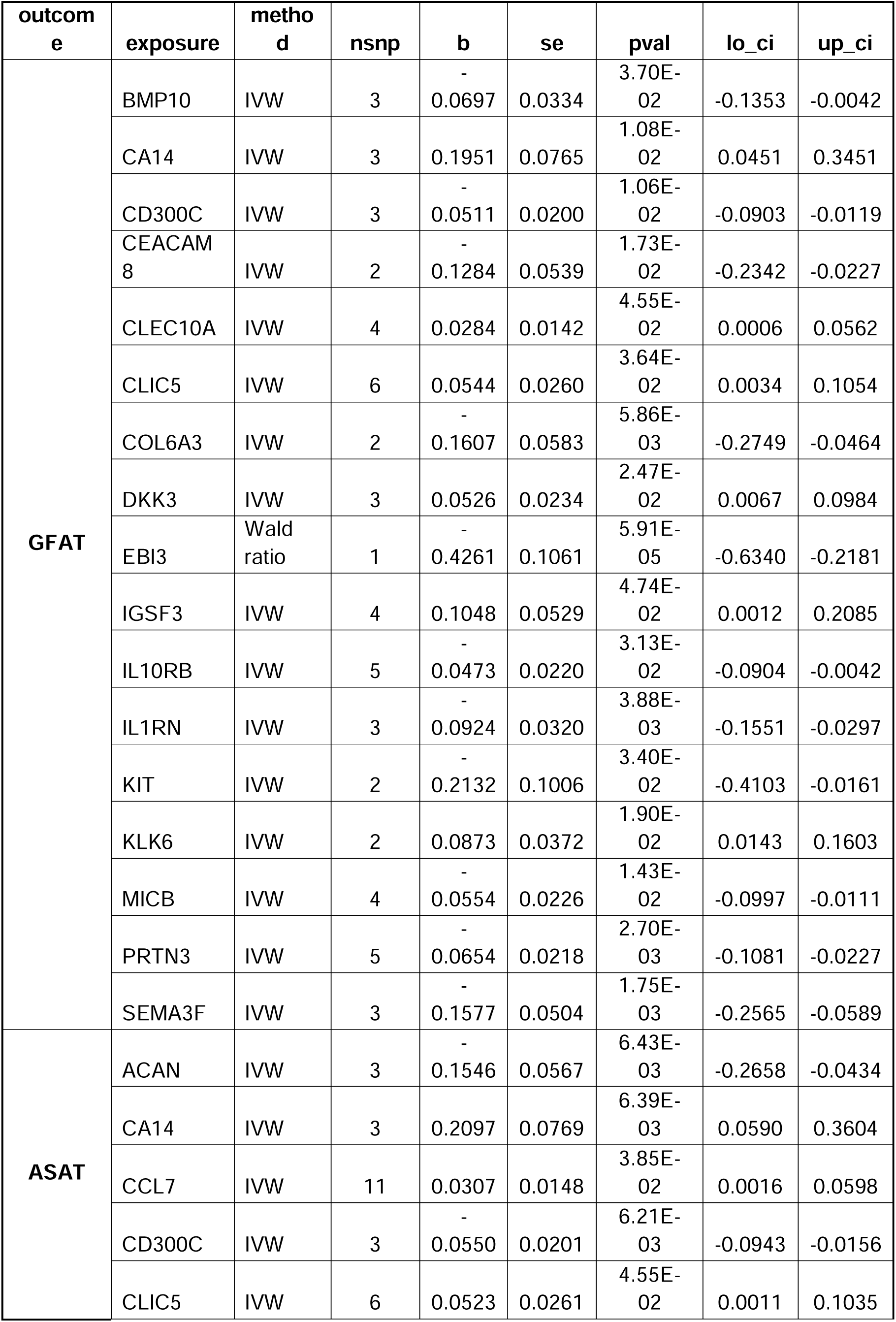
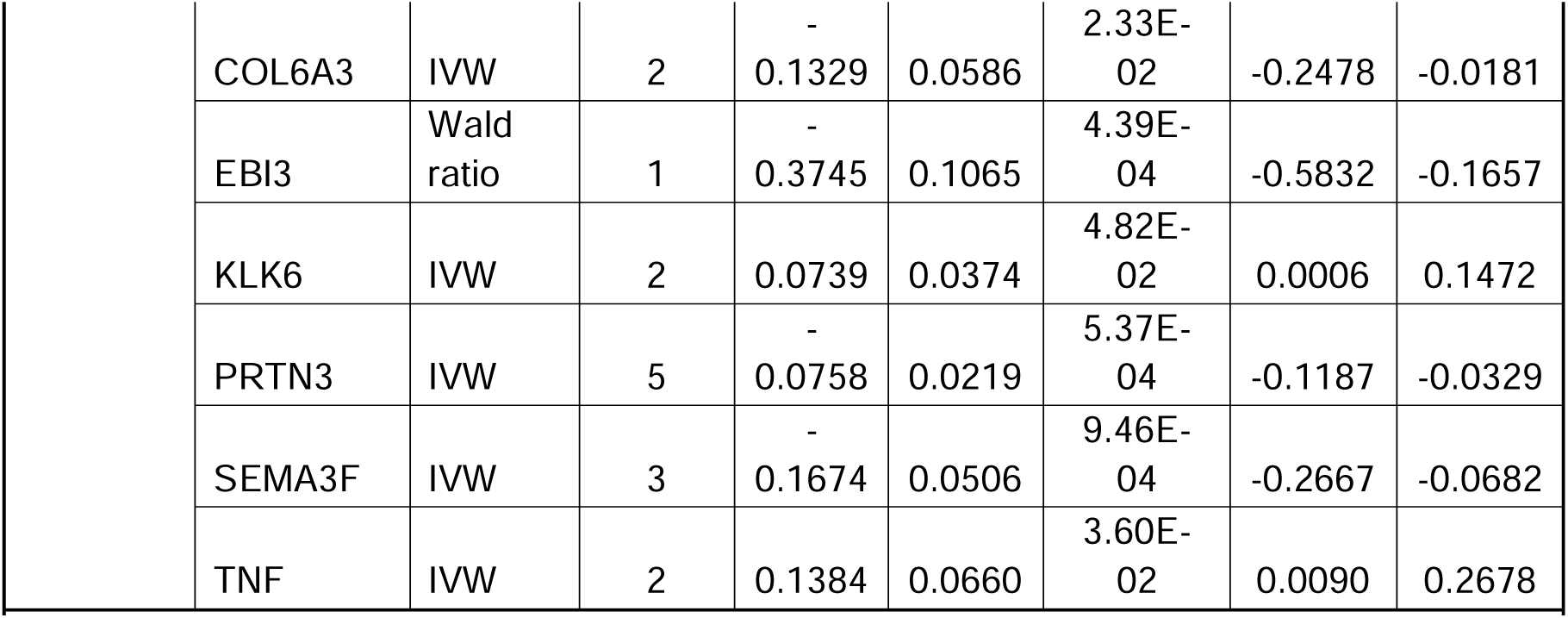
Two-step MR: IVW or Wald ratio effects for candidate pQTL effects on ASAT and GFAT (Step 2). Summary of results for candidate pQTLs that are both influenced by cBMI during the temporal window of interest, and demonstrate evidence of an effect on ASAT or GFAT.

In contrast, several proteins showed positive associations with ASAT, including CA14 (beta= 0.21, 95% CI= 0.06, 0.36) and TNF (beta= 0.14, 95% CI= 0.01, 0.27), while KLK6 (beta= 0.07, 95% CI= 0.00, 0.15), CLIC5 (beta= 0.05, 95% CI= 0.00, 0.10), and CCL7 were also positively associated, albeit with a smaller effect size (beta= 0.03, 95% CI= 0.00, 0.06)

For GFAT, 17 candidate proteins showed evidence consistent with a causal effect (**Table 3**). Several proteins showed inverse associations with GFAT, including EBI3 (beta= −0.43, 95% CI= −0.63, −0.22), KIT (beta= −0.21, 95% CI= −0.41, −0.02), SEMA3F (beta= −0.16, 95% CI= −0.26 to −0.06), COL6A3 (beta= −0.16, 95% CI= −0.28, −0.05), and CEACAM8 (beta= −0.13, 95% CI= −0.23, −0.02). Inverse associations were also observed for CD300C, MICB, IL1RN, IL10RB, PRTN3, and BMP10.

Conversely, several proteins were positively associated with GFAT. CA14 (beta= 0.20, 95% CI= 0.05, 0.35), IGSF3 (beta= 0.10, 95% CI= 0.00, 0.21), KLK6 (beta= 0.09, 95% CI= 0.01, 0.16), and CLIC5 (beta= 0.05, 95% CI= 0.00, 0.11) showed positive effects. Modest positive associations were also observed for DKK3 and CLEC10A.

Together, these results identify a refined subset of circulating proteins that may mediate the effect of childhood BMI on adult fat distribution, with eleven candidate proteins identified for ASAT and seventeen for GFAT, of which 8 proteins appear as candidates for both (CA14, CD300C, CLIC5, COL6A3, EBI3, KLK6, PRNT3, SEMA3F).

### Summary of two-step MR

The final stage of the analysis focused on identifying candidate mediators that were directionally consistent across the full causal sequence from cBMI to protein levels and subsequently to adiposity outcomes. Given that higher cBMI was associated with increased ASAT and GFAT, only proteins for which the direction of effect in Step 1 aligned with the direction of effect in Step 2 were retained. Directional consistency was defined as either (i) cBMI increasing protein levels and higher protein levels in turn increasing adiposity, or (ii) cBMI decreasing protein levels and higher protein levels decreasing adiposity, implying that reduced protein levels would be associated with increased adiposity. Proteins showing opposing directions across steps were excluded, as these patterns were inconsistent with the observed cBMI–adiposity relationships (**Table 4**).

**Table 4.**
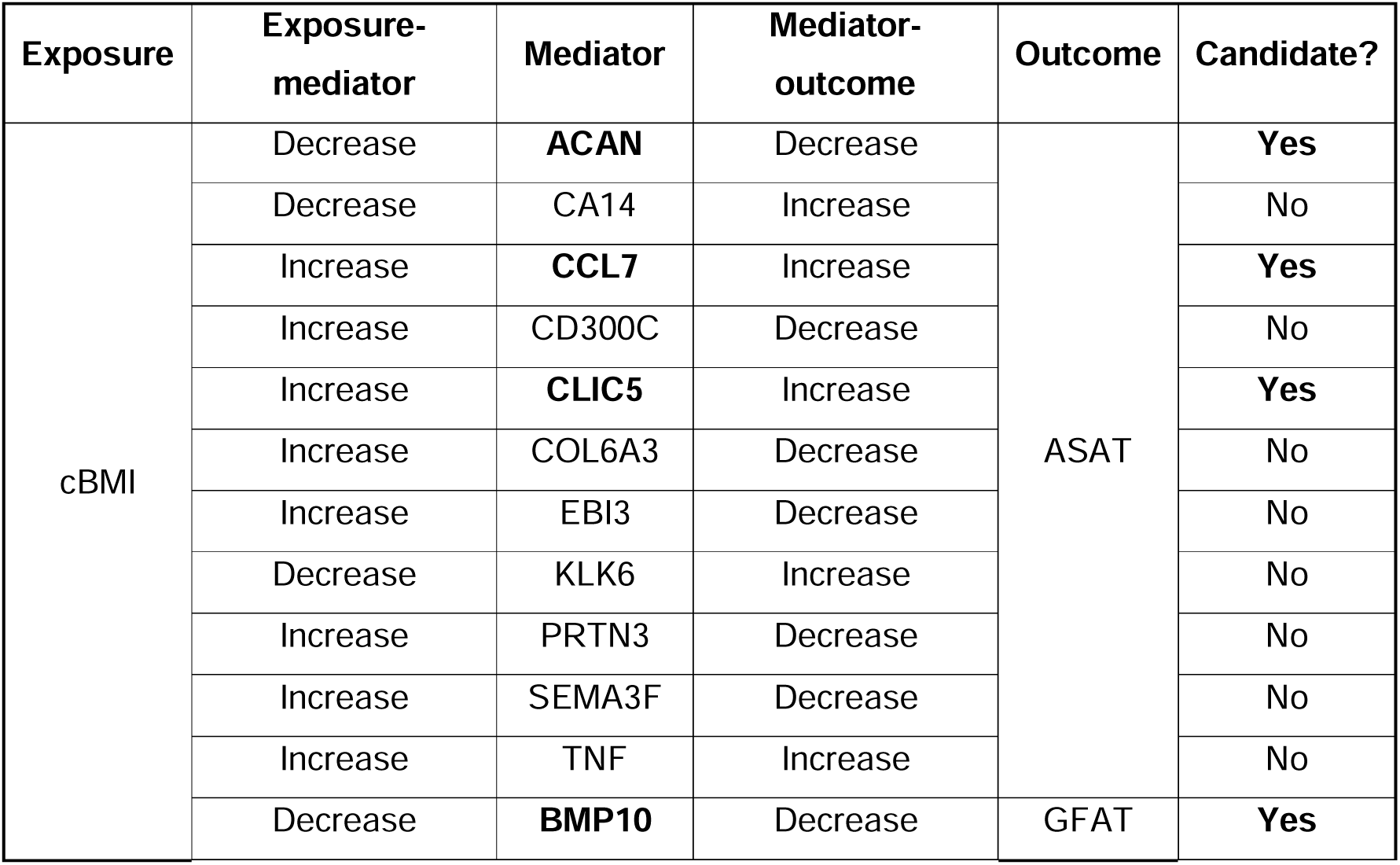
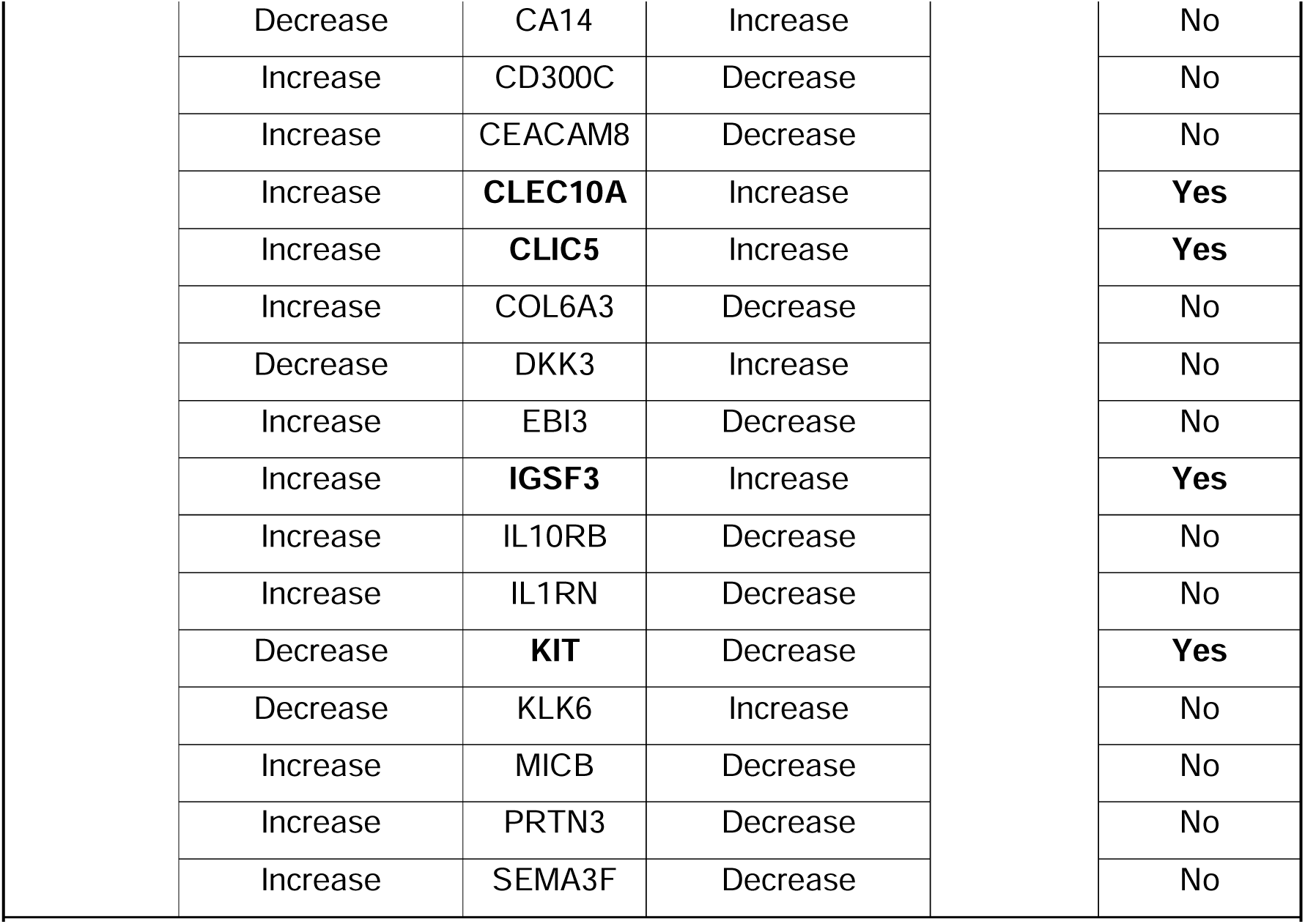
Summary of direction of effects for two-step MR. Exposure-mediator (step 1) refers to cBMI effect on protein levels, mediator-outcome (step 2) refers to protein level effect on ASAT and GFAT volume. Analyses that are directionally consistent with the primary MR results across all steps are considered candiate mediators and taken forward for further analysis.

Applying temporal and directional consistency criteria resulted in three candidate mediators for ASAT and five for GFAT, with one candidate overlapping between the two (CLIC5) (**Figure 5**).

For ASAT, these included ACAN, for which cBMI was associated with decreased protein levels and higher ACAN levels were associated with lower ASAT, indicating that cBMI-related reductions in ACAN may contribute to increased ASAT. Two additional proteins, CLIC5 and CCL7, had directionally consistent effects, with cBMI-associated increases in protein levels corresponding to increased ASAT.

For GFAT, three proteins met the criteria for directional consistency across both MR steps. These included CLEC10A, CLIC5, and IGSF3. In each case, the direction of the cBMI–protein association in Step 1 was concordant with the direction of the protein–GFAT association in Step 2, consistent with the observed effect of cBMI on GFAT accumulation. Additionally, cBMI was associated with decreased levels of BMP10 and KIT, both of which were associated with lower GFAT, indicating that cBMI-related reductions in these proteins may contribute to increased GFAT. All remaining candidate proteins were excluded due to discordant directions of effect between the two MR steps.

As a supplementary analysis, reverse two-sample MR was conducted (**Supplementary Table 13**). Evidence of reverse causation was observed for a small subset of proteins, primarily in relation to ASAT. Specifically, higher ASAT was associated with lower circulating ACAN levels (beta= −0.14, 95% CI= −0.27, −0.01), indicating a potential bidirectional relationship. In addition, a positive association with ASAT were observed for CCL7 (beta= 0.19, 95% CI= 0.05, 0.32). These findings suggest that these proteins may act as both mediators and downstream consequences of ASAT, and causal inference for these candidates should therefore be interpreted with caution. No evidence of reverse causation was observed for CLIC5 in relation to ASAT, supporting its role as a potential mediator of the effect of cBMI on ASAT.

In contrast, CLIC5 showed borderline evidence of a reciprocal association with GFAT. We also found evidence of a positive association between GFAT and CLEC10A, indicating that both of these proteins may act as both a mediator and confounder in the childhood BMI–GFAT pathway. No evidence of reverse effects was observed for BMP10, KIT, or IGSF3, supporting their interpretation as potential mediators of the effect of cBMI on GFAT.

Steiger filtering was also conducted for protein effects on ASAT and GFAT, but none of the exposure SNPs were found to be more strongly associated with the fat depot outcomes. This represents an alternative approach to inferring causal direction, though this method carries its own limitations, particularly sensitivity to differences in statistical power between the discovery GWAS for exposure and outcome.

Overall, this sequential two-step MR approach substantially reduced the number of plausible mediating proteins by integrating temporal, statistical, and directional evidence. The final set of seven candidate mediators represents proteins most likely to lie on the causal pathway linking childhood BMI to adult fat distribution.

### Exploration of candidate proteins

Having identified ACAN, CCL7 and CLIC5 as candidate mediators of childhood BMI effects on ASAT, and CLIC5, BMP10, CLEC10A, KIT and IGSF3 as candidate mediators of childhood BMI effects on GFAT, we sought to characterise the mechanistic basis through which these proteins may influence fat depot volume. We first tested whether the genetic signals underlying each candidate protein and the respective fat depot GWAS shared a common causal variant, using pairwise conditional and colocalisation analysis (PWCoCo). Across all eight protein-fat depot pairs, colocalisation analysis consistently returned high posterior probabilities for H1 (range: 0.92–0.93), with negligible support for a shared causal variant (H4 range: 0.018–0.019). This pattern indicates that the genetic signal in each tested region was specific to the protein trait, with little evidence of a corresponding association with the fat depot outcome. (**Supplementary Table 14**). This suggests that the candidate proteins act upstream in a causal chain rather than sharing a common genetic architecture with ASAT or GFAT at the tested loci, and motivated further investigation into potential intermediate mechanisms.

To assess whether the genetic variants instrumenting circulating levels of each candidate protein also regulate gene expression in metabolically relevant tissue - consistent with the direction of effect in our MR framework - we queried the GTEx eQTL Dashboard (v8) using each protein’s lead pQTL SNP and its corresponding gene, with subcutaneous adipose as the tissue of interest, given the strength of evidence found for ASAT and GFAT depots (**Table 5**). The lead pQTL SNPs for BMP10 (rs34008398, p=0.008) and CLEC10A (rs90951, p=0.002) were significantly associated with expression of their respective genes in subcutaneous adipose tissue, indicating that the genetic signal instrumenting these circulating proteins also influences local gene expression in adipose, consistent with a role for adipose tissue in mediating downstream effects on fat depot volume. CLIC5 showed a marginal eQTL association (rs2022423, p=0.059). No evidence of adipose eQTL effects was observed for ACAN, CCL7, KIT, or IGSF3, suggesting that for these proteins, the relevant tissue context may lie elsewhere, or that regulation operates through post-transcriptional or systemic mechanisms not captured by adipose eQTL analysis.

**Table 5.** GTEx eQTL effects of candidate protein pQTL variants in subcutaneous adipose tissue. Lead pQTL SNPs for each candidate protein were queried against the GTEx v10 eQTL Dashboard (Adipose – Subcutaneous). Median refers to average gene expression by genotype group (homozygous reference, heterozygous, and homozygous alternative allele carriers). Significance was defined as p<0.05

| Gene | SNP | Homozygous ref |  | Heterozygous |  | Homozygous alt |  | pval | Significant? |
| --- | --- | --- | --- | --- | --- | --- | --- | --- | --- |
|  |  | N | median | N | median | N | median |  |  |
| CLIC5 | rs2022423 | 285 | 0.01056 | 339 | 0.0176 | 87 | -0.1913 | 5.88E-02 | Marginal |
| BMP10 | rs34008398 | 696 | -0.0088 | 15 | 0.5513 | 0 | NA | 8.37E-03 | Yes |
| CLEC10A | rs90951 | 261 | 0.03874 | 310 | -0.04931 | 140 | 0.0546 | 2.08E-03 | Yes |
| KIT | rs78744187 | 623 | -0.00704 | 81 | 0.04578 | 7 | -0.05636 | 8.98E-01 | No |
| IGSF3 | rs56278466 | 71 | -0.1591 | 301 | -0.01408 | 339 | 0.03169 | 3.62E-01 | No |
| CCL7 | rs12075 | 108 | 0.07047 | 339 | 0.0176 | 264 | 0.09697 | 2.90E-01 | No |
| ACAN | rs1245582 | 91 | 0.1377 | 336 | -0.01056 | 284 | -0.01584 | 1.99E-01 | No |

To identify traits associating with each candidate protein that may help to understand their biological context and potential relevance to fat depot outcomes, we performed a phenome-wide association study (PheWAS) for each protein, restricting analyses to continuous traits, risk factors, and metabolites (approximately 5,700 outcomes per protein after exclusion of ancestry-mismatched GWAS) and applying both Bonferroni and Benjamini-Hochberg FDR correction (**Supplementary table 15)**. Of the eight candidate proteins, only KIT yielded associations surviving multiple testing correction. Genetically predicted KIT levels were robustly associated with white blood cell count (β=−0.316, p=6.77×10⁻ ³³), neutrophil count (β=−0.412, p=1.46×10⁻³¹), neutrophil percentage (β=−0.343, p=1.38×10⁻ ²³), and lymphocyte percentage (β=0.372, p=4.42×10⁻²&#x25A1;), all surviving Bonferroni correction. Monocyte count (β=−0.201, p=1.61×10⁻ &#x25A1;) and a brain imaging phenotype reflecting functional connectivity in resting-state network 100 component 603 (NET100 0603; β=−0.935, p=4.23×10⁻ &#x25A1;) additionally passed FDR correction (**Table 6**).

**Table 6.**
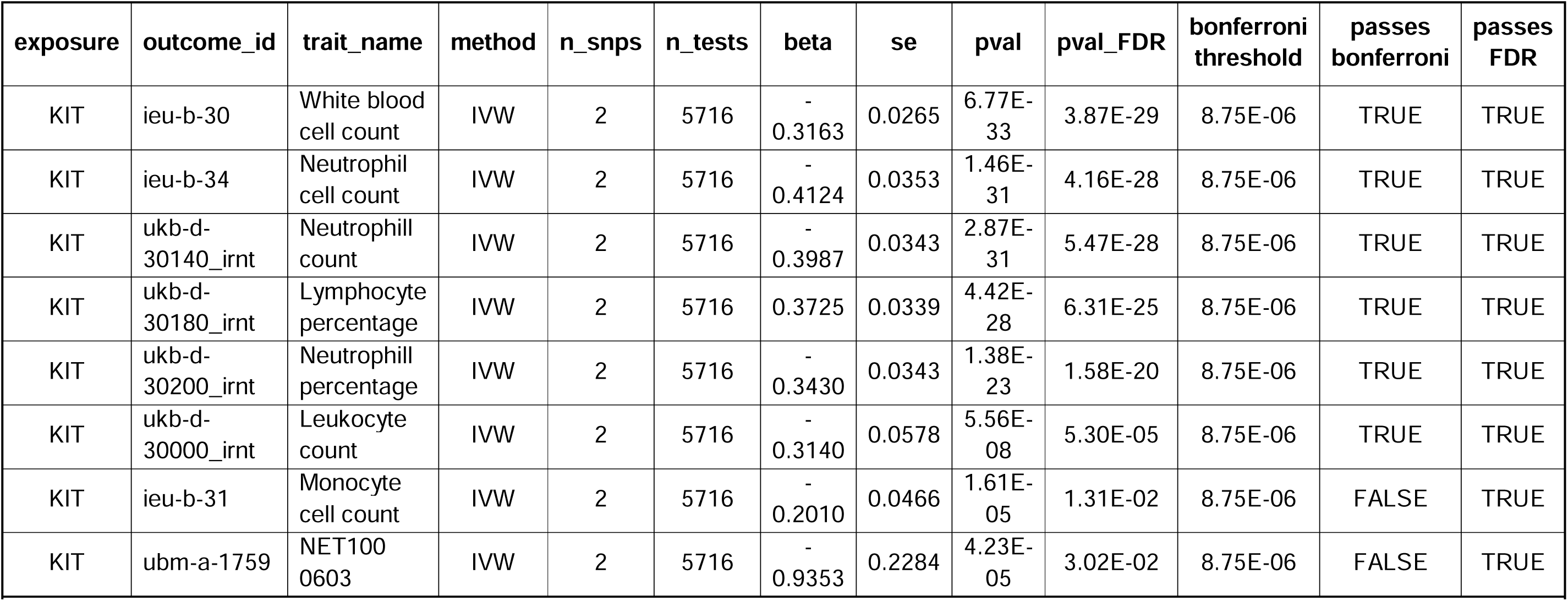
PheWAS hits for proteins surviving FDR correction. Results are shown for genetically predicted KIT levels as exposure, restricted to outcomes passing FDR correction from a PheWAS of approximately 5,700 ancestry-matched outcomes from the IEU OpenGWAS database. Beta represents the SD change in outcome per SD change in genetically predicted KIT levels. Bonferroni threshold was calculated based on the number of unique outcomes tested for KIT following exclusion of ancestry-mismatched GWAS (n=5,716)

| exposure | outcome_id | trait_name | method | n_snps | n_tests | beta | se | pval | pval_FDR | bonferroni threshold | passes bonferroni | passes FDR |
| --- | --- | --- | --- | --- | --- | --- | --- | --- | --- | --- | --- | --- |
| KIT | ieu-b-30 | White blood cell count | IVW | 2 | 5716 | -0.3163 | 0.0265 | 6.77E-33 | 3.87E-29 | 8.75E-06 | TRUE | TRUE |
| KIT | ieu-b-34 | Neutrophil cell count | IVW | 2 | 5716 | -0.4124 | 0.0353 | 1.46E-31 | 4.16E-28 | 8.75E-06 | TRUE | TRUE |
| KIT | ukb-d-30140_irnt | Neutrophill count | IVW | 2 | 5716 | -0.3987 | 0.0343 | 2.87E-31 | 5.47E-28 | 8.75E-06 | TRUE | TRUE |
| KIT | ukb-d-30180_irnt | Lymphocyte percentage | IVW | 2 | 5716 | 0.3725 | 0.0339 | 4.42E-28 | 6.31E-25 | 8.75E-06 | TRUE | TRUE |
| KIT | ukb-d-30200_irnt | Neutrophill percentage | IVW | 2 | 5716 | -0.3430 | 0.0343 | 1.38E-23 | 1.58E-20 | 8.75E-06 | TRUE | TRUE |
| KIT | ukb-d-30000_irnt | Leukocyte count | IVW | 2 | 5716 | -0.3140 | 0.0578 | 5.56E-08 | 5.30E-05 | 8.75E-06 | TRUE | TRUE |
| KIT | ieu-b-31 | Monocyte cell count | IVW | 2 | 5716 | -0.2010 | 0.0466 | 1.61E-05 | 1.31E-02 | 8.75E-06 | FALSE | TRUE |
| KIT | ubm-a-1759 | NET100 0603 | IVW | 2 | 5716 | -0.9353 | 0.2284 | 4.23E-05 | 3.02E-02 | 8.75E-06 | FALSE | TRUE |

## DISCUSSION

In this study, we used an MR framework to investigate the causal effects of cBMI on adult fat distribution and to identify circulating proteins that may mediate these effects. We found that higher genetically predicted cBMI, from age 7 years onwards, was causally associated with increased ASAT and GFAT depots in adulthood. In contrast, there was little evidence of causal effects on VAT, liver fat, or pancreatic fat, suggesting that the influence of childhood adiposity on adult body composition is depot-specific rather than generalised. Replication of the direction and timing of effects for subcutaneous depots using stronger instruments from the original discovery GWAS further reinforces the validity of the main IVW findings and supports the interpretation that childhood BMI exerts a more pronounced influence on adult subcutaneous fat distribution than on ectopic or visceral depots.

Building on these findings, two-step MR analyses identified evidence for a subset of circulating proteins whose levels were influenced by childhood BMI during a developmentally sensitive window - characterised here as the period spanning three to twelve years of age, at which point cBMI effects on ASAT and GFAT first emerged - and which in turn exerted causal effects on ASAT and/or GFAT. By integrating temporal consistency, depot specificity, and directional concordance across MR steps, we identified three candidate mediators for ASAT and five for GFAT. Together, these results support the existence of biologically distinct and temporally sensitive pathways linking childhood adiposity to adult fat distribution. Notably, the predominant pattern across the 140 candidate proteins was one of cBMI acquiring influence over the circulating proteome as development progresses, with 136 proteins classified as gain-of-effect candidates, suggesting that mid-childhood represents a period of increasing susceptibility to adiposity-related molecular change.

### Comparison with other studies

Our findings align with previous epidemiological and genetic studies reporting that higher BMI in childhood is associated with greater adult adiposity [48], particularly in subcutaneous depots. Observational cohorts have shown that adiposity accrued during childhood and adolescence continues into adulthood, with stronger associations for subcutaneous rather than visceral fat [49]. Genetic studies using adult BMI instruments have similarly reported depot-specific effects [31]. While the importance of adiposity timing across the life course and developmental factors such as pubertal timing have been explored in relation to later disease and cardiometabolic outcomes[50,51], we are not aware of studies specifically examining how childhood adiposity at different developmental stages influences adult fat distribution. Moreover, to our knowledge, no previous study has integrated age-specific genetic instruments with two-step MR to investigate circulating proteins as potential mediators of these relationships. Our findings therefore extend existing work by incorporating both temporal resolution and molecular mediation to identify developmentally sensitive pathways linking childhood adiposity with adult fat distribution.

Several MR studies investigating early-life adiposity have suggested that childhood BMI exerts long-lasting effects on adult metabolic traits, including cardiometabolic risk factors, with some associations appearing to persist independently of adult BMI [52,53]. However, evidence from life-course multivariable MR analyses has suggested that this pattern may not apply universally across outcomes. For example, studies of cardiovascular disease outcomes accounting for adult body size have shown substantial attenuation of associations, indicating that the influence of childhood adiposity may often be mediated indirectly through adult adiposity rather than acting independently across all traits [54]. Our results build on this literature by suggesting that these long-term effects are not uniformly distributed across fat depots and appear instead to preferentially affect ASAT and GFAT. This depot specificity is consistent with known differences in the developmental origin, cellular composition, and metabolic function of adipose tissue across anatomical sites, and suggests that the pathways linking early-life adiposity to adult body composition are more nuanced than a generalised effect on fat accumulation.

The emergence of effects around mid-childhood (age 7 years) is consistent with developmental studies indicating that adipose tissue expansion and differentiation accelerate during this period, potentially increasing susceptibility to lasting structural and metabolic changes [55,56]. The consistency in direction and temporal pattern across sex-stratified and combined analyses further supports the robustness of these findings, with sex-specific differences appearing to reflect differences in magnitude rather than the presence or absence of an effect.

With respect to molecular mediation, previous proteomic studies have identified numerous proteins associated with BMI and adiposity in cross-sectional analyses [57,58]. Our findings extend this literature by providing evidence for specific circulating proteins as plausible causal mediators in the developmental pathway from childhood BMI to adult fat depot volume. The candidate proteins identified span several biological processes relevant to adipose tissue biology. For ASAT, ACAN — a large extracellular matrix proteoglycan — was identified as a candidate mediator, with cBMI-associated reductions in ACAN potentially contributing to increased ASAT through altered ECM scaffolding and adipose tissue development [59]. CCL7, a CC chemokine involved in monocyte and macrophage recruitment [60], and CLIC5, a chloride intracellular channel protein implicated in cytoskeletal organisation and membrane integrity [61], were also identified as ASAT mediators, consistent with a role for immune and cytoskeletal signalling in subcutaneous adipose accumulation. For GFAT, candidate mediators included CLIC5, BMP10 - a bone morphogenetic protein involved in vascular and developmental signalling [62] - CLEC10A, a C-type lectin receptor expressed on antigen-presenting cells [63], KIT, a receptor tyrosine kinase critical for haematopoiesis (hence findings from PheWAS) [48], and IGSF3, an immunoglobulin superfamily member involved in cell adhesion [64]. The partially distinct protein profiles for ASAT and GFAT highlight biological heterogeneity in the mechanisms linking childhood BMI to adult fat distribution and are consistent with the known differences in immune cell composition and developmental origin between subcutaneous abdominal and gluteofemoral depots.

Follow-up analyses of these candidate proteins provided further mechanistic insight. Colocalisation analyses indicated that pQTL signals for all candidate proteins were independent of the ASAT and GFAT GWAS signals, consistent with the proteins acting upstream rather than at a shared locus. GTEx eQTL analyses revealed that the cis pQTL variants for BMP10 and CLEC10A also regulate gene expression in subcutaneous adipose tissue, implicating this tissue as a site of action for these proteins. For KIT, PheWAS analyses identified robust associations with circulating white blood cell, neutrophil, and lymphocyte measures, raising the possibility that KIT influences GFAT through immune-mediated mechanisms, given the recognised role of adipose tissue immune infiltration in depot-specific fat accumulation [49]. The biological relevance of the NET100 0603 brain imaging association observed for KIT is less clear and may reflects pleiotropy at the KIT locus rather than a meaningful mechanistic link to GFAT. The absence of PheWAS hits for the remaining proteins suggests that their mechanistic pathways may operate through traits not captured in the current phenome scan, or that instrument power was insufficient to detect associations across the broader phenome.

### Strengths and limitations of this study

Key strengths of this study include the use of genetically informed causal inference methods, longitudinally modelled childhood BMI instruments spanning early childhood to late adolescence, and imaging-derived measures of adult fat distribution for defining depot-specific outcomes. The two-step MR framework allowed us to integration of temporal, statistical, and directional evidence, thereby reducing the risk of spurious mediator identification.

Several limitations should also be considered. First, instrument strength for childhood BMI varied across timepoints, with weaker instruments at younger ages. This likely reflects, at least in part, the greater phenotypic variability in BMI at older ages - as growth trajectories diverge across individuals during late childhood and adolescence. As a result, genetic instruments may capture variation more reliably at older ages, irrespective of underlying causal effects. This variation in precision may have reduced power to detect early-life effects and contributed to greater uncertainty in sensitivity analyses such as MR-Egger, which is particularly sensitive to weak instrument bias. However, replication using stronger MoBa instruments supported the timing of observed effects, suggesting findings are unlikely to be driven entirely by weak instrument bias.

A further interpretational limitation is that we were not able to disentangle direct effects of childhood BMI from those mediated through adult adiposity using multivariable Mendelian randomisation (MVMR). This was not feasible due unavailability of full childhood BMI summary statistics, limited instrument strength for age-specific BMI measures, and high correlation between genetic predictors across developmental stages, which can lead to weak instrument bias in MVMR analyses. As a result, the childhood BMI estimates may partly reflect pathways operating through later-life adiposity.

A related limitation concerns the construction of childhood BMI genetic instruments across developmental stages. Although the replication instruments were derived from BMI measured in early childhood (birth to age 8 years), they were applied across a wider developmental window in ALSPAC (ages 3 to 18 years). This assumes partial stability in the genetic architecture of BMI across childhood and adolescence, although age-specific genetic effects and changing environmental influences may lead to some heterogeneity in instrument validity over time. As such, age-specific effect estimates should be interpreted with appropriate caution.

The use of circulating protein levels may also represent a limitation, as these may not fully capture tissue-specific expression or local effects within adipose depots, potentially underestimating the role of some mediators. Integration with external resources such as GTEx suggests that some candidate proteins show heterogeneous expression across tissues, supporting the possibility that circulating measures may underestimate depot-specific biological effects. In addition, evidence of bidirectional effects for certain proteins, particularly ACAN and CCL7 in relation to ASAT, suggests that some candidates may act as both mediators and downstream consequences of adiposity, complicating causal interpretation for these proteins specifically.

A further structural limitation of our analysis is the potential for sample overlap between the GWAS used to derive pQTL instruments (UKB-PPP) and the imaging-derived GWAS for ASAT and GFAT, both of which draw on overlapping subsets of UK Biobank participants. In two-sample MR, sample overlap can introduce bias towards observational associations, particularly when instruments are weak [34,38]. Although the pQTL instruments used here were generally strong (selected at genome-wide significance and LD-clumped), and thus less susceptible to weak instrument bias amplification, the degree of overlap cannot be precisely quantified. We therefore cannot fully exclude some bias in protein–fat depot estimates, and these findings should be interpreted with appropriate caution. This limitation does not apply to the primary childhood BMI analyses, which used non-overlapping cohorts for instrument derivation and outcome assessment.

Finally, analyses were restricted to individuals of predominantly European ancestry, limiting generalisability to more diverse populations. This is particularly relevant given known differences in BMI distribution, genetic architecture, and cardiometabolic risk across ancestries. Prior work has also highlighted disparities in representation of non-European populations in both genetic studies and clinical research, underscoring the need for greater diversity in study cohorts and downstream validation of findings [65]. Future work in more ancestrally diverse cohorts will therefore be important to assess external validity and ensure applicability beyond European-ancestry populations. In addition, the PheWAS was restricted to continuous traits, risk factors, and metabolites, meaning that additional mechanistic pathways operating through other phenotypic domains may have been missed.

### Implications of findings

These findings have several important implications. First, they suggest that the timing of adiposity accumulation in childhood is critical, with mid-childhood representing a potential window during which BMI exerts lasting effects on adult fat distribution. This reinforces the importance of early-life interventions aimed at preventing excessive weight gain before adolescence. While pharmacological interventions for obesity, including Glucagon-Like Peptide-1 (GLP-1) receptor agonists, have recently been approved for use in adolescents aged 12 years and older [66], our findings suggest that earlier life stages may represent a period of heightened susceptibility in which prevention may be more impactful than treatment once obesity is established.

Second, the identification of depot-specific mediating proteins highlights biological heterogeneity in the pathways linking childhood BMI to adult adiposity, suggesting that abdominal and gluteofemoral fat may be influenced by overlapping but distinct molecular mechanisms, with implications for understanding the differential metabolic consequences of these depots. Third, the refined list of candidate mediators provides a focused set of targets for future functional studies, which may help elucidate mechanisms of adipose tissue development and identify potential biomarkers or intervention targets.

More broadly, this work exemplifies the utility of combining lifecourse MR with multi-omics data to interrogate developmental origins of adult body composition. The analytical framework applied here could be extended to other early-life exposures and downstream metabolic outcomes.

## Conclusion

In conclusion, this study provides suggestive causative evidence that higher childhood BMI from approximately age 7 years onwards increases adult subcutaneous fat accumulation, particularly in abdominal and gluteofemoral depots. Using a two-step MR approach, we identified seven circulating proteins that may mediate these effects, with partially distinct mediating profiles for ASAT and GFAT. These findings advance understanding of the biological pathways linking early-life adiposity to adult fat distribution and underscore the importance of childhood as a critical period for shaping long-term adipose tissue patterns.

## METHODS

### Study Design and Objectives

This study is reported in accordance with the STROBE-MR (Strengthening the Reporting of Observational Studies in Epidemiology using Mendelian Randomisation) guidelines [25]. A completed STROBE-MR checklist is provided in **Supplementary document 1**.

We conducted a Mendelian randomisation (MR) study to investigate whether childhood body mass index (cBMI) has a causal effect on the distribution of body fat in adulthood, and to identify candidate proteins that may mediate this effect. Firstly, we conducted two-sample MR to explore the effect of cBMI at 12 timepoints between the ages of 3 and 18 years on regional fat deposition in adults (mean age 64.5 years (SD 7.7 years)), including ASAT, GFAT, VAT, liver fat, and pancreas fat. Only those fat depots found to be influenced by cBMI in primary MR analyses were taken forward for mediation analyses (see Results; **Figure 1**).

Subsequently, we implemented a two-step MR framework to investigate mediating effects [24]. In Step 1, we assessed the effects of cBMI across the lifecourse on 2,940 circulating proteins measured using Olink proteomics. In Step 2, we evaluated whether candidate proteins exert causal effects on cBMI–associated fat depots.

### Genetic Instruments for Childhood BMI

We obtained publicly-available genetic instruments for childhood BMI from the Norwegian Mother, Father and Child Cohort Study (MoBa), a large population-based pregnancy cohort that has prospectively followed families in Norway since 1999, comprising up to 28,681 children with repeated BMI measurements across early development [26]. BMI was measured at 12 time points from birth to 8 years of age (birth, 6 weeks, 3, 6 and 8 months; and 1, 1.5, 2, 3, 5, 7 and 8 years; **Supplementary Table 1**). At each timepoint, standardised BMI was analysed using linear mixed-effects models under an additive genetic model. This resulted in 46 independent SNPs reaching genome-wide significance (r^2^=0.2, PO<O5O×O10⁻ O) at one or more timepoints [27].

For replication and primary effect estimation, data from the Avon Longitudinal Study of Parents and Children (ALSPAC) were used (N=736 - 5780). ALSPAC is a UK-based prospective birth cohort that recruited over 14,000 pregnant women between 1991 and 1992, which aimed to investigate the genetic and environmental determinants of health and development in parents and children [28,29]. Using instruments from the MoBa discovery GWAS, childhood BMI was modelled across a series of timepoints in ALSPAC [27], with those replicating at P<0.05 taken forward to model cBMI at ages 3, 5, 6, 7, 8, 10, 12, 14, 15, 16, 17, and 18 years in the primary analyses of this study (**Supplementary Table 1)**. ALSPAC-derived effect estimates were used in preference to those from MoBa for two reasons. First, using discovery sample effect estimates in MR risks winner’s curse bias, whereby SNP-trait associations are overestimated due to instrument selection in the same sample, potentially biasing downstream causal estimates [30]. Second, ALSPAC provided timepoints spanning ages 3–18 years compared to birth to 8 years of age from MoBa, enabling investigation over a longer time period for the time course MR analyses. Although F-statistics derived from ALSPAC effect estimates were lower than those from MoBa (Fstat = ∼5–11 vs. ∼33–35), the potential for weak instrument bias was assessed through sensitivity analyses and considered explicitly in the interpretation of results.

### Genetic Instruments for Circulating Proteins (pQTLs)

Genetic instruments for circulating protein levels were obtained from genome-wide association studies of plasma proteins measured using the antibody-based Olink Explore 3072 platform in UK Biobank participants (N= 54,219) [23]. For each of the proteins identified as candidates in step-1 of the two-step MR, genetic instruments were identified using a P-value threshold of 5 x 10^-8^.

To minimise bias from linkage disequilibrium and horizontal pleiotropy, only independent variants (r² < 0.01 within a 1 Mb window) were retained for each protein. All pQTL instruments were harmonised to the fat depot outcome datasets prior to analysis. Full details of selected pQTL instruments are provided in **Supplementary Table 2**.

### Outcome Data: Fat Distribution in Adulthood

We used published genome-wide association study (GWAS) summary statistics for five MRI-derived measures of fat distribution from the UK Biobank: Abdominal subcutaneous, gluteofemoral, and visceral fat (N= 38,964) [31]; and liver (N= 32,858) and pancreas fat (N= 25617) [32]. Each outcome was adjusted for ten genetic principal components, age, sex, body size, and imaging covariates, and transformed to approximate normality.

### Primary MR Analyses: Childhood BMI to Adult Fat Distribution

We conducted two-sample MR to estimate the effect of cBMI (at 12 childhood timepoints) on adult fat distribution. These analyses were used to identify both the fat depots influenced by cBMI and the developmental window during which effects emerged, thereby informing subsequent two-step MR analyses (Results; **Figure 1)**. For all main analyses, any results with a p-value below 0.05 were classed as having evidence for an effect, but were also subject to further scrutiny using a series of sensitivity analyses to assess robustness. To account for multiple testing, both Benjamini-Hochberg false discovery rate (FDR) correction and Bonferroni correction were applied [33], with results interpreted in light of both thresholds alongside the nominal p-value.

### Two-step MR Analyses: Step 1

We conducted two-sample MR to estimate the effects of cBMI at each timepoint on adult circulating protein levels. Protein quantitative trait loci (pQTLs) for 2,940 Olink-measured proteins were used as outcomes in Step 1 [23].

### Two-step MR Analyses: Step 2

In the second step of the two-step MR framework, candidate proteins identified in Step 1 were evaluated for causal effects on adult fat depots. Only adiposity outcomes that demonstrated evidence of a causal effect of cBMI in the primary MR analyses were considered in this step.

Two-sample MR analyses were conducted using genome-wide significant (P<5×10^-8^) genetic instruments for each candidate protein as exposures and fat depots as outcomes.

### Statistical and sensitivity analyses

MR relies on three core assumptions: (i) relevance, that genetic instruments are robustly associated with the exposure; (ii) independence, that instruments are not associated with confounders of the exposure–outcome relationship; and (iii) exclusion restriction, that instruments affect the outcome only through the exposure. Instrument strength (relevance) was assessed using F-statistics, while the independence and exclusion restriction assumptions were evaluated using a series of pleiotropy-robust sensitivity analyses, described below.

The primary MR method for all analyses was the inverse variance weighted (IVW) estimator, under the assumption that all genetic instruments are valid or that any horizontal pleiotropy is balanced [34]. To assess the robustness of IVW estimates to potential pleiotropy, several complementary MR methods were applied, including MR-Egger regression, weighted median, simple mode, and weighted mode approaches [35–37]. Where only a single genetic instrument was available, Wald ratio estimates were calculated.

Instrument strength was assessed using F-statistics, calculated for each exposure–outcome analysis. Mean F-statistics were reported to evaluate the potential for weak instrument bias, with values below 10 indicating limited instrument strength [38]. For MR-Egger analyses, SIMEX correction was applied to account for regression dilution bias arising from weak instruments [39].

Additional sensitivity analyses included MR-LASSO, which identifies and down-weights potentially invalid instruments [40]; MR-PRESSO, which detects horizontal pleiotropic outliers and provides outlier-corrected estimates [41]; and Radial MR, which facilitates identification of influential variants and assesses heterogeneity before and after outlier removal [42]. Cochran’s Q statistic was used to quantify heterogeneity across instruments.

Reverse two-sample MR was conducted to assess whether fat depots exert causal effects on the candidate mediating proteins. Evidence of effects in the reverse direction may suggest bidirectional relationships; however, the interpretation of reverse MR findings is not straightforward, as the presence of a reverse effect does not necessarily negate a causal effect in the forward direction [43]. Results of the reverse MR should therefore be interpreted cautiously and considered alongside the primary analyses rather than as definitive evidence against forward causality.

Sex-stratified MR analyses to explore the effects of cBMI and PQTLs on male- and female-specific depot outcomes were conducted to further assess robustness. All analyses were performed in R (version 4.2.0) using the “TwoSampleMR”, “RadialMR”, “MRPRESSO”, and “MendelianRandomization” packages.

### Candidate Mediator Selection

Candidate mediators taken forward into Step 2 were identified using a multi-stage filtering strategy. First, only those proteins where there was evidence for a causal effect (P<0.05) of cBMI at any timepoint on protein levels were taken forward.

Proteins showing no evidence of association (P>0.05) were excluded. Second, proteins were required to demonstrate temporally consistent effects of cBMI on protein levels across the developmental window identified in the primary analyses (defined as 3 time points either side of where effect of cBMI on fat depot first emerged). Candidate mediating proteins were therefore required to mirror the pattern identified in the primary analysis — that is, to show a consistent presence or absence of effect across timepoints, rather than sporadic associations. For example, a protein showing an effect of cBMI at age 7, no effect at ages 8–9, a further effect at age 10, and no effect at ages 12 or 14 would be considered inconsistent and excluded. Conversely, proteins demonstrating consistent causal effects across all timepoints were also excluded, as these are unlikely to capture mechanisms underlying an age-specific shift in fat deposition.

Proteins meeting these criteria were classified as either gain of effect (GOE) or loss of effect (LOE) candidates. GOE candidates were proteins where cBMI showed no effect prior to the developmental window (e.g., up to cBMI at age 7 years) and a persistent effect thereafter (e.g., from age 7 years onwards), whereas LOE candidates were proteins where cBMI showed evidence of effect up to the developmental window followed by no effect at later timepoints.

### Colocalisation of candidate proteins with fat depots

To determine whether the genetic signals underlying circulating levels of each candidate protein and the respective fat depot GWAS shared a common causal variant, which is necessary for the two traits to have a causal relationship, we performed pairwise conditional and colocalisation analysis using PWCoCo [44]. Colocalisation was performed within a ±500kb window around the lead pQTL SNP for each protein, using prior probabilities p1=5×10⁻&#x25A1;, p2=5×10⁻ &#x25A1; and p12=1×10⁻&#x25A1;, consistent with recommended priors for cis-pQTL colocalisation analyses. Posterior probabilities were extracted for each colocalisation hypothesis: H0 (no association with either trait), H1 (association with the protein signal only), H2 (association with the cancer outcome only), H3 (both traits associated but driven by distinct variants), and H4 (both traits associated and sharing a single causal variant). Evidence of colocalisation was defined by H4.

### GTEx eQTL analysis

To assess whether the genetic variants instrumenting circulating levels of each candidate protein also regulate gene expression in depot-relevant adipose tissue - consistent with the direction of effect in our MR framework - we queried the GTEx eQTL Dashboard (v10) using each protein’s lead pQTL SNP and corresponding

Ensembl gene identifier, with “adipose” selected as the tissue of interest. Results were considered significant at p<0.05 after inspection of the full p-value distribution across all gene-variant pairs.

### Phenome-wide association study (PheWAS)

To identify traits associated with both candidate protein levels and fat depot measures, without inferring directionality, we conducted a PheWAS for each candidate protein using the IEU OpenGWAS database as the outcome source [45,46]. Available GWAS were filtered to three categories — Continuous, Risk factor, and Metabolites — using the OpenGWAS annotation catalogue, yielding approximately 5,700 ancestry-matched outcomes per protein. Ancestry-mismatched GWAS (identified by outcome ID suffixes indicating non-European ancestry, including UKB East Asian and BioBank Japan datasets) were excluded prior to analysis, as pQTL instruments were derived from European-ancestry samples. MR analyses were performed using the TwoSampleMR package in R, applying the Wald ratio method for single-instrument exposures and inverse variance weighted (IVW) meta-analysis where multiple instruments were available. Multiple testing was addressed by calculating Bonferroni-corrected thresholds based on the number of unique outcomes tested per protein following ancestry exclusion, and by applying Benjamini-Hochberg false discovery rate (FDR) correction within each protein’s PheWAS [33]. IVW results derived from fewer than five SNPs were flagged as potentially unreliable.

## Data Availability

All data are available online:
Summary statistics for liver fat and pancreas fat GWAS are available from the NHGRI-EBI GWAS Catalog, accession numbers GCST90016673 (liver fat) and GCST90016675 (pancreas fat). http://ftp.ebi.ac.uk/pub/databases/gwas/summary_statistics/GCST90016001-GCST90017000/GCST90016676/).
GWAS summary statistics for ASAT, GFAT and VAT are available for download from the Cardiovascular Disease Knowledge Portal under the "Downloads" tab (https://cvd.hugeamp.org/).
Genetic instruments for childhood BMI were obtained from the supplementary data of Helgeland et al. (2022), Nature Metabolism (https://doi.org/10.1038/s42255-022-00549-1).
Proteomic GWAS (pQTL) summary statistics from the UK Biobank Pharma Proteomics Project (UKB-PPP) are available via the Synapse platform (https://www.synapse.org/Synapse:syn51364943), with pGWAS summary statistics specifically available at https://www.synapse.org/Synapse:syn51365301.

https://cvd.hugeamp.org/

https://doi.org/10.1038/s42255-022-00549-1

https://www.synapse.org/Synapse:syn51365301

## List of abbreviations

ASAT: Abdominal subcutaneous adipose tissue
ALSPAC: Avon Longitudinal Study of Parents and Children
BMI: Body mass index
cBMI: Childhood body mass index
eQTL: Expression quantitative trait loci
FDR: False discovery rate
GFAT: Gluteofemoral adipose tissue
GLP-1: Glucagon-like-peptide 1
GOE: Gain of effect
GWAS: Genome wide association study
IVW: Inverse variance weighted
LOE: Loss of effect
MOBA: Norwegian Mother, Father and Child Cohort Study
MR: Mendelian randomisation
MVMR: Multivariable Mendelian randomisation
pQTL: Protein quantitative trait loci
SD: Standard deviation
SNP: Single nucleotide polymorphism
UKB-PPP: UK Biobank Pharma Proteomics Project
VAT: Visceral adipose tissue

## Declarations

### Ethics approval and consent to participate

This study is an analysis of publicly available, summary-level genetic association data and did not involve the collection of new data, recruitment of participants, or access to individual-level data. No separate ethical approval was required.

Ethical approval and participant consent for the original studies contributing data were obtained as follows. For ALSPAC, ethical approval was obtained from the ALSPAC Ethics and Law Committee (IRB00003312) and the Local Research Ethics Committees, with informed consent for the use of data collected via questionnaires and clinics obtained from participants in accordance with ALSPAC Ethics and Law Committee recommendations. For MoBa, the establishment of the cohort and initial data collection were based on a license from the Norwegian Data Protection Agency and approval from the Regional Committees for Medical and Health Research Ethics, Norway. For UK Biobank and the UK Biobank Pharma Proteomics Project (UKB-PPP), ethical approval was obtained from the North West Multi-Centre Research Ethics Committee (Ref: 11/NW/0382).

### Consent for publication

Not applicable

### Availability of data and materials

Summary statistics for liver fat and pancreas fat GWAS are available from the NHGRI-EBI GWAS Catalog, accession numbers GCST90016673 (liver fat) and GCST90016675 (pancreas fat). http://ftp.ebi.ac.uk/pub/databases/gwas/summary_statistics/GCST90016001-GCST90017000/GCST90016676/).

GWAS summary statistics for ASAT, GFAT and VAT are available for download from the Cardiovascular Disease Knowledge Portal under the “Downloads” tab (https://cvd.hugeamp.org/).

Genetic instruments for childhood BMI were obtained from the supplementary data of Helgeland et al. (2022), Nature Metabolism (https://doi.org/10.1038/s42255-022-00549-1).

Proteomic GWAS (pQTL) summary statistics from the UK Biobank Pharma Proteomics Project (UKB-PPP) are available via the Synapse platform (https://www.synapse.org/Synapse:syn51364943), with pGWAS summary statistics specifically available at https://www.synapse.org/Synapse:syn51365301.

### Disclaimer

Where authors are identified as personnel of the International Agency for Research on Cancer/World Health Organization, the authors alone are responsible for the views expressed in this article, and they do not necessarily represent the decisions, policy, or views of the International Agency for Research on Cancer/World Health Organization.

### Authors’ contributions

BH conceived and designed the study, performed the statistical analysis, and drafted the manuscript. RR and EV supervised the project, contributed to the conception and design of the study, and helped shape the structure and interpretation of the manuscript. All other authors critically reviewed and edited the manuscript. All authors read and approved the final manuscript.

## Acknowledgements

Not applicable

## Figure Legends

**Figure 1. Primary MR analysis: Childhood BMI effect on adult fat deposition.**

Two-sample MR to demonstrate effect of childhood BMI (ages 3-18 years) on adult fat deposition (ASAT, GFAT, VAT, and liver and pancreatic fat). Effect estimates in early childhood were generally modest, with stronger and more consistent associations emerging from mid-childhood at age 7 years onwards. 2a shows results as a forest plot, and 2b shows results graphically.

**Figure 2. MR analysis: Step 1; Childhood BMI effect on proteins**

Two-sample MR to demonstrate effect of childhood BMI (ages 3-18 years) on circulating protein levels for 136 “gain of effect” and 4 “loss of effect” candidate mediators.

**Figure 3. MR analysis: Step 2; pQTL effect on ASAT and GFAT.**

Two-sample MR to demonstrate effect of circulating protein levels for 140 candidate mediators, resulting in 11 significant candidates for 4a) ASAT; and 17 significant candidates for 4b) GFAT.\

**Figure 4. Summary of two-step MR analysis.**

Childhood BMI effect on candidate pQTLs during temporal window of interest (step 1) and pQTL effect on ASAT and GFAT (step 2). Blue arrows indicate a decrease in outcome, red arrows indicate an increase in outcome. Green arrows indicate evidence of a confounding effect.

**Figure 5. Study design overview.**

Childhood BMI (ages 3-18 years) was used as the exposure in both the primary MR analyses to explore effects on adult fat distribution (ASAT, GFAT, VAT, and liver and pancreatic fat), and also in Step 1 of two-step MR to explore effects on circulating protein levels. Step 2 shows circulating protein levels assessed as potential mediators, which were then filtered for temporal and directional consistency, before undergoing mechanistic analyses.

## References

1. Zhang X, Liu J, Ni Y, Yi C, Fang Y, Ning Q, et al. Global Prevalence of Overweight and Obesity in Children and Adolescents: A Systematic Review and Meta-Analysis. JAMA pediatrics. 2024 Aug;178(8):800–13.

2. Abarca-Gómez L, Abdeen ZA, Hamid ZA, Abu-Rmeileh NM, Acosta-Cazares B, Acuin C, et al. Worldwide trends in body-mass index, underweight, overweight, and obesity from 1975 to 2016: a pooled analysis of 2416 population-based measurement studies in 128&#xb7;9 million children, adolescents, and adults. The Lancet. 2017 Dec 16;390(10113):2627–42.

3. Kerr JA, Patton GC, Cini KI, Abate YH, Abbas N, Abd Al Magied AHA, et al. Global, regional, and national prevalence of child and adolescent overweight and obesity, 1990-2021, with forecasts to 2050: a forecasting study for the Global Burden of Disease Study 2021. The Lancet. 2025 Mar 8;405(10481):785–812.

4. Wei Y, Richardson TG, Zhan Y, Carlsson S. Childhood adiposity and novel subtypes of adult-onset diabetes: a Mendelian randomisation and genome-wide genetic correlation study. Diabetologia. 2023 Jun;66(6):1052–6.

5. Umer A, Kelley GA, Cottrell LE, Giacobbi PJ, Innes KE, Lilly CL. Childhood obesity and adult cardiovascular disease risk factors: a systematic review with meta-analysis. BMC public health. 2017 Aug;17(1):683.

6. Mandic M, Safizadeh F, Schöttker B, Hoffmeister M, Brenner H. Association of childhood-to-adulthood body size change with cancer risk: UK Biobank prospective cohort. BMC medicine. 2025 May;23(1):268.

7. Hazelwood E, Goudswaard LJ, Lee MA, Vabistsevits M, Pournaras DJ, Brenner H, et al. Adiposity distribution and risks of 12 obesity-related cancers: a Mendelian randomization analysis. JNCI: Journal of the National Cancer Institute. 2025 Dec 1;117(12):2621–42.

8. Lee M-J, Wu Y, Fried SK. Adipose tissue heterogeneity: implication of depot differences in adipose tissue for obesity complications. Molecular aspects of medicine. 2013 Feb;34(1):1–11.

9. Luo L, Liu M. Adipose tissue in control of metabolism. The Journal of endocrinology. 2016 Dec;231(3):R77–99.

10. An S-M, Cho S-H, Yoon JC. Adipose Tissue and Metabolic Health. Diabetes & metabolism journal. 2023 Sep;47(5):595–611.

11. Murphy N, Newton CC, Song M, Papadimitriou N, Hoffmeister M, Phipps AI, et al. Body mass index and molecular subtypes of colorectal cancer. Journal of the National Cancer Institute. 2023 Feb;115(2):165–73.

12. Wu M, Huang Y, Liu Q. Relationship between body mass index and cardiovascular metabolic multimorbidity: a systematic review and meta-analysis. Frontiers in cardiovascular medicine. 2025;12:1568348.

13. Kivimäki M, Strandberg T, Pentti J, Nyberg ST, Frank P, Jokela M, et al. Body-mass index and risk of obesity-related complex multimorbidity: an observational multicohort study. The lancet Diabetes & endocrinology. 2022 Apr;10(4):253–63.

14. Khan SS, Ning H, Wilkins JT, Allen N, Carnethon M, Berry JD, et al. Association of Body Mass Index With Lifetime Risk of Cardiovascular Disease and Compression of Morbidity. JAMA cardiology. 2018 Apr;3(4):280–7.

15. Fontvieille E, Viallon V, Recalde M, Cordova R, Jansana A, Peruchet-Noray L, et al. Body mass index and cancer risk among adults with and without cardiometabolic diseases: evidence from the EPIC and UK Biobank prospective cohort studies. BMC medicine. 2023 Nov;21(1):418.

16. Gupta A, Pandey A, Ayers C, Beg MS, Lakoski SG, Vega GL, et al. An Analysis of Individual Body Fat Depots and Risk of Developing Cancer: Insights From the Dallas Heart Study. Mayo Clinic proceedings. 2017 Apr;92(4):536–43.

17. Lu Y, Zhao YC, Liu K, Bever A, Zhou Z, Wang K, et al. A validated estimate of visceral adipose tissue volume in relation to cancer risk. Journal of the National Cancer Institute. 2024 Dec;116(12):1942–51.

18. Rubino F, Cummings DE, Eckel RH, Cohen R V, Wilding JPH, Brown WA, et al. Definition and diagnostic criteria of clinical obesity. The Lancet Diabetes & Endocrinology. 2025 Mar 1;13(3):221–62.

19. Hammerton G, Munafò MR. Causal inference with observational data: the need for triangulation of evidence. Psychological medicine. 2021 Mar;51(4):563–78.

20. Davey Smith G, Hemani G. Mendelian randomization: genetic anchors for causal inference in epidemiological studies. Human molecular genetics. 2014/07/04. 2014 Sep 15;23(R1):R89–98.

21. Davies NM, Holmes M V., Davey Smith G. Reading Mendelian randomisation studies: A guide, glossary, and checklist for clinicians. BMJ (Online). 2018;362.

22. Smith GD, Ebrahim S. “Mendelian randomization”: Can genetic epidemiology contribute to understanding environmental determinants of disease? International Journal of Epidemiology. 2003;32(1):1–22.

23. Sun BB, Chiou J, Traylor M, Benner C, Hsu Y-H, Richardson TG, et al. Plasma proteomic associations with genetics and health in the UK Biobank. Nature. 2023 Oct;622(7982):329–38.

24. Relton CL, Davey Smith G. Two-step epigenetic Mendelian randomization: a strategy for establishing the causal role of epigenetic processes in pathways to disease. International journal of epidemiology. 2012 Feb;41(1):161–76.

25. Skrivankova VW, Richmond RC, Woolf BAR, Davies NM, Swanson SA, Vanderweele TJ, et al. Strengthening the reporting of observational studies in epidemiology using mendelian randomisation (STROBE-MR): Explanation and elaboration. The BMJ. 2021;375.

26. Magnus P, Birke C, Vejrup K, Haugan A, Alsaker E, Daltveit AK, et al. Cohort Profile Update: The Norwegian Mother and Child Cohort Study (MoBa). International Journal of Epidemiology. 2016 Apr 1;45(2):382–8.

27. Helgeland Ø, Vaudel M, Sole-Navais P, Flatley C, Juodakis J, Bacelis J, et al. Characterization of the genetic architecture of infant and early childhood body mass index. Nature metabolism. 2022 Mar;4(3):344–58.

28. Boyd A, Golding J, Macleod J, Lawlor DA, Fraser A, Henderson J, et al. Cohort Profile: the ‘children of the 90s’--the index offspring of the Avon Longitudinal Study of Parents and Children. International journal of epidemiology. 2013 Feb;42(1):111–27.

29. Fraser A, Macdonald-Wallis C, Tilling K, Boyd A, Golding J, Davey Smith G, et al. Cohort Profile: the Avon Longitudinal Study of Parents and Children: ALSPAC mothers cohort. International journal of epidemiology. 2013 Feb;42(1):97–110.

30. Bowden J, Dudbridge F. Unbiased estimation of odds ratios: combining genomewide association scans with replication studies. Genetic Epidemiology. 2009 Jul 1;33(5):406–18.

31. Agrawal S, Wang M, Klarqvist MDR, Smith K, Shin J, Dashti H, et al. Inherited basis of visceral, abdominal subcutaneous and gluteofemoral fat depots. Nature communications. 2022 Jun;13(1):3771.

32. Liu Y, Basty N, Whitcher B, Bell JD, Sorokin EP, van Bruggen N, et al. Genetic architecture of 11 organ traits derived from abdominal MRI using deep learning. eLife. 2021 Jun;10.

33. Benjamini Y, Hochberg Y. Controlling the False Discovery Rate: A Practical and Powerful Approach to Multiple Testing. Journal of the Royal Statistical Society Series B (Methodological). 1995 Jun 1;57(1):289–300.

34. Burgess S, Butterworth A, Thompson SG. Mendelian randomization analysis with multiple genetic variants using summarized data. Genetic epidemiology. 2013 Nov;37(7):658–65.

35. Hartwig FP, Davey Smith G, Bowden J. Robust inference in summary data Mendelian randomization via the zero modal pleiotropy assumption. International journal of epidemiology. 2017 Dec 1;46(6):1985–98.

36. Bowden J, Smith GD, Burgess S. Mendelian randomization with invalid instruments: Effect estimation and bias detection through Egger regression. International Journal of Epidemiology. 2015;44(2):512–25.

37. Bowden J, Davey Smith G, Haycock PC, Burgess S. Consistent Estimation in Mendelian Randomization with Some Invalid Instruments Using a Weighted Median Estimator. Genetic epidemiology. 2016 May;40(4):304–14.

38. Burgess S, Thompson SG. Avoiding bias from weak instruments in mendelian randomization studies. International Journal of Epidemiology. 2011;40(3):755–64.

39. Hardin JW, Schmiediche H, Carroll RJ. The Simulation Extrapolation Method for Fitting Generalized Linear Models with Additive Measurement Error. The Stata Journal: Promoting communications on statistics and Stata. 2003;3(4):373–85.

40. Rees JMB, Wood AM, Dudbridge F, Burgess S. Robust methods in Mendelian randomization via penalization of heterogeneous causal estimates. PLOS ONE. 2019 Sep 23;14(9):e0222362.

41. Verbanck M, Chen CY, Neale B, Do R. Detection of widespread horizontal pleiotropy in causal relationships inferred from Mendelian randomization between complex traits and diseases. Nature Genetics. 2018;50(5):693–8.

42. Bowden J, Spiller W, Del Greco M F, Sheehan N, Thompson J, Minelli C, et al. Improving the visualization, interpretation and analysis of two-sample summary data Mendelian randomization via the Radial plot and Radial regression. International journal of epidemiology. 2018 Aug;47(4):1264–78.

43. Bull C, Hazelwood E, Bell JA, Tan V, Constantinescu A-E, Borges C, et al. Identifying metabolic features of colorectal cancer liability using Mendelian randomization. Hägg S, Franco EL, editors. eLife. 2023;12:RP87894.

44. Robinson JW, Hemani G, Babaei MS, Huang Y, Baird DA, Tsai EA, et al. An efficient and robust tool for colocalisation: Pair-wise Conditional and Colocalisation (PWCoCo). bioRxiv. 2022 Jan 1;2022.08.08.503158.

45. Denny JC, Bastarache L, Roden DM. Phenome-Wide Association Studies as a Tool to Advance Precision Medicine. Annual review of genomics and human genetics. 2016 Aug;17:353–73.

46. Hemani G, Elsworth B, Palmer T, Rasteiro R. ieugwasr: Interface to the “OpenGWAS” Database API. 2025.

47. Khera A V, Chaffin M, Wade KH, Zahid S, Brancale J, Xia R, et al. Polygenic Prediction of Weight and Obesity Trajectories from Birth to Adulthood. Cell. 2019 Apr;177(3):587–596.e9.

48. Guo SS, Huang C, Maynard LM, Demerath E, Towne B, Chumlea WC, et al. Body mass index during childhood, adolescence and young adulthood in relation to adult overweight and adiposity: the Fels Longitudinal Study. International journal of obesity and related metabolic disordersÖ: journal of the International Association for the Study of Obesity. 2000 Dec;24(12):1628–35.

49. Kindblom JM, Lorentzon M, Hellqvist A, Lönn L, Brandberg J, Nilsson S, et al. BMI changes during childhood and adolescence as predictors of amount of adult subcutaneous and visceral adipose tissue in men: the GOOD Study. Diabetes. 2009 Apr;58(4):867–74.

50. Bell JA, Carslake D, Wade KH, Richmond RC, Langdon RJ, Vincent EE, et al. Influence of puberty timing on adiposity and cardiometabolic traits: A Mendelian randomisation study. PLoS medicine. 2018 Aug;15(8):e1002641.

51. Harroud A, Mitchell RE, Richardson TG, Morris JA, Forgetta V, Davey Smith G, et al. Childhood obesity and multiple sclerosis: A Mendelian randomization study. Multiple sclerosis (Houndmills, Basingstoke, England). 2021 Dec;27(14):2150–8.

52. Richardson TG, Sanderson E, Elsworth B, Tilling K, Davey Smith G. Use of genetic variation to separate the effects of early and later life adiposity on disease risk: mendelian randomisation study. BMJ. 2020 May 6;369:m1203.

53. Richardson TG, Mykkänen J, Pahkala K, Ala-Korpela M, Bell JA, Taylor K, et al. Evaluating the direct effects of childhood adiposity on adult systemic metabolism: a multivariable Mendelian randomization analysis. International journal of epidemiology. 2021 Nov;50(5):1580–92.

54. Power GM, Tyrrell J, Frayling TM, Davey Smith G, Richardson TG. Mendelian Randomization Analyses Suggest Childhood Body Size Indirectly Influences End Points From Across the Cardiovascular Disease Spectrum Through Adult Body Size. Journal of the American Heart Association. 2021 Sep;10(17):e021503.

55. Palacios-Marin I, Serra D, Jiménez-Chillarón JC, Herrero L, Todorčević M. Childhood obesity: Implications on adipose tissue dynamics and metabolic health. Obesity Reviews. 2023 Dec 1;24(12):e13627.

56. Orsso CE, Colin-Ramirez E, Field CJ, Madsen KL, Prado CM, Haqq AM. Adipose Tissue Development and Expansion from the Womb to Adolescence: An Overview. Nutrients. 2020 Sep;12(9).

57. Avocegamou R, Jumentier B, Fagbemi K, Yazdanpanah N, Yazdanpanah M, Gamache I, et al. Proteome-wide mendelian randomization reveals circulating proteins causally associated with childhood body mass index. Scientific Reports. 2025;16(1):2093.

58. Goudswaard LJ, Bell JA, Hughes DA, Corbin LJ, Walter K, Davey Smith G, et al. Effects of adiposity on the human plasma proteome: observational and Mendelian randomisation estimates. International journal of obesity (2005). 2021 Oct;45(10):2221–9.

59. Voros G, Sandy JD, Collen D, Lijnen HR. Expression of aggrecan(ases) during murine preadipocyte differentiation and adipose tissue development. Biochimica et biophysica acta. 2006 Dec;1760(12):1837–44.

60. Chang T-T, Chen C, Chen J-W. CCL7 as a novel inflammatory mediator in cardiovascular disease, diabetes mellitus, and kidney disease. Cardiovascular diabetology. 2022 Sep;21(1):185.

61. Jiang L, Phang JM, Yu J, Harrop SJ, Sokolova A V, Duff AP, et al. CLIC proteins, ezrin, radixin, moesin and the coupling of membranes to the actin cytoskeleton: A smoking gun? Biochimica et Biophysica Acta (BBA) - Biomembranes. 2014;1838(2):643–57.

62. Desroches-Castan A, Tillet E, Bouvard C, Bailly S. BMP9 and BMP10: Two close vascular quiescence partners that stand out. Developmental Dynamics. 2022 Jan 1;251(1):158–77.

63. Qin Y, Wang L, Zhang L, Li J, Liao L, Huang L, et al. I mmunological role and prognostic potential of CLEC10A in pan-cancer. American journal of translational research. 2022;14(5):2844–60.

64. Guo Y, Kasai Y, Tanaka Y, Ohashi-Kumagai Y, Sakamoto T, I to T, et al. IGSF3 is a homophilic cell adhesion molecule that drives lung metastasis of melanoma by promoting adhesion to vascular endothelium. Cancer science. 2024 Jun;115(6):1936–47.

65. Alsaqaaby MS, Cooney S, le Roux CW, Pournaras DJ. Sex, race, and BMI in clinical trials of medications for obesity over the past three decades: a systematic review. The Lancet Diabetes & Endocrinology. 2024;12(6):414–21.

66. Salama M, Hassan D, Kumar S. Updates on Anti-Obesity Medications in Children and Adolescents. Children (Basel, Switzerland). 2025 Oct;12(10).

