## Supplementary document 1 for "Causal Effects of Childhood BMI on Regional Fat Distribution in Adults: A Mendelian Randomisation and Proteomic Mediation Analysis"

### STROBE-MR Checklist

*Page numbers omitted — to be added before submission. Rows shaded orange mark items not addressed in the manuscript text reviewed and needing text added; rows shaded yellow are partially addressed and may need a short addition.*

| **Item** | **Section** | **Checklist item** | **Page number** | **Relevant text from manuscript** |
| --- | --- | --- | --- | --- |
| 1 | TITLE and ABSTRACT | Indicate MR as the study's design in the title and/or abstract. | 0-1 | Causal Effects of Childhood BMI on Regional Fat Distribution in Adults: A Mendelian Randomisation and Proteomic Mediation Analysis |
| **INTRODUCTION** | | | | |
| 2 | Background | Explain the scientific background and rationale. What is the exposure? Is a causal relationship plausible? Why is MR helpful? | 2-4 | Background section establishes childhood obesity as a growing global concern, explains the rationale for examining depot-specific fat distribution rather than BMI alone, and states that “observational analyses are often susceptible to confounding and reverse causality, limiting the ability to infer causality”, before introducing MR as a method that “offers a robust tool to address these limitations… provided that strong assumptions hold”. |
| 3 | Objectives | State specific objectives clearly, including pre-specified hypotheses. State that MR estimates causal effects under specific assumptions. | 3-4 | Final paragraph of Background: “we use MR to investigate the causal effect of childhood BMI (12 timepoints from 3 to 18 years) on adult fat distribution measures, specifically ASAT, GFAT, VAT, liver fat, and pancreas fat… we systematically assess the role of circulating proteins in mediating these relationships by analysing 2,940 plasma proteins… using a two-step MR approach.”  Explanation of MR assumptions already addressed previously. |
| **METHODS** | | | | |
| 4a | Setting | Describe study design, underlying population, setting, locations, relevant dates. | 5 | Methods describes: MoBa (population-based pregnancy cohort, Norway, recruiting since 1999); ALSPAC (UK birth cohort, Bristol, recruiting >14,000 pregnant women 1991–1992); UK Biobank MRI substudy for fat depot outcomes (mean age 64.5 years, SD 7.7); UKB-PPP for proteomic data (Olink Explore 3072 platform). |
| 4b | Participants | Give eligibility criteria and sources/methods of participant selection. Report sample size and any power calculations. | 5-6 | cBMI instruments: MoBa discovery GWAS (up to 28,681 children); ALSPAC replication/primary analysis, N=736–5,780 across 12 timepoints. Fat depot outcome GWAS (UK Biobank): ASAT/GFAT/VAT N=38,964; liver fat N=32,858; pancreas fat N=25,617. Proteomic GWAS (UKB-PPP): N=54,219. |
| 4c |  | Describe measurement, quality control and selection of genetic variants. | 5-6 | cBMI: standardised BMI modelled with linear mixed-effects models under an additive genetic model at 12 MoBa timepoints, yielding 46 independent SNPs (r²=0.2, P<5×10⁻⁸). SNPs replicated in MoBa at P<0.05. pQTL instruments: selected at P<5×10⁻⁸ and LD-clumped (r²<0.01, 1 Mb window); harmonised to outcome datasets prior to analysis. |
| 4d |  | For each exposure, outcome and other variable, describe methods of assessment and diagnostic criteria. | 5-6 | cBMI assessed at 12 age-specific timepoints (birth–18 years) via standardised BMI in MoBa/ALSPAC. Adult fat depots: MRI-derived ASAT, GFAT, VAT, liver and pancreas fat in UK Biobank, adjusted for 10 genetic PCs, age, sex, body size and imaging covariates, transformed to approximate normality. Circulating proteins: 2,940 plasma proteins quantified via the Olink Explore 3072 platform (UKB-PPP). |
| 4e |  | Provide details of ethics committee approval and participant informed consent, if relevant. | 27 | Covered in the Declarations / “Ethics approval and consent to participate” section rather than in the Methods. |
| 5 | Assumptions | Explicitly state the three core IV assumptions (relevance, independence, exclusion restriction) for the main analysis, and assumptions for sensitivity analyses. | 7-9 | Covered in the “statistical and sensitivity analyses” section of the methods. “MR relies on three core assumptions: (i) relevance, that genetic instruments are robustly associated with the exposure; (ii) independence, that instruments are not associated with confounders of the exposure–outcome relationship; and (iii) exclusion restriction, that instruments affect the outcome only through the exposure…” |
| 6a | Statistical methods: main analysis | Describe how quantitative variables were handled (scale, units, model). | 7-9 | Effect estimates reported as change in MRI-derived fat depot volume (SD units) per 1 kg/m² increase in genetically predicted cBMI; protein–depot associations reported as SD change in fat depot volume per SD increase in genetically predicted protein level. |
| 6b |  | Describe how genetic variants were handled and, if applicable, how weights were selected. | 7-9 | Independent variants retained after LD-clumping (r²<0.01, 1 Mb window) for protein instruments; cBMI instruments selected at r²=0.2, P<5×10⁻⁸ across the 12 timepoints; all instruments harmonised to outcome datasets prior to analysis. |
| 6c |  | Describe the MR estimator and related statistics; covariates used, and whether the same covariate set was used in both samples for two-sample MR. | 7-9 | IVW used as the primary estimator (assuming instrument validity or balanced pleiotropy); Wald ratio used for single-instrument exposures. Outcome GWAS (UK Biobank fat depots) adjusted for 10 genetic PCs, age, sex, body size and imaging covariates.. |
| 6d |  | Explain how missing data were addressed. | 7-9 | Where exposure SNPs were not available in the outcome data, proxy SNPs were identified using the harmonise_data() function in the TwosampleMR package. |
| 6e |  | If applicable, indicate how multiple testing was addressed. | 7 | Both Benjamini–Hochberg FDR correction and Bonferroni correction applied throughout (candidate mediator selection and PheWAS), with results interpreted against both thresholds alongside nominal p-values. |
| 7 | Assessment of assumptions | Describe methods or prior knowledge used to assess the assumptions or justify their validity. | 7-9 | Instrument strength assessed via F-statistics; robustness to pleiotropy (exclusion-restriction violations) assessed via MR-Egger, weighted median and mode-based estimators; colocalisation (PWCoCo) used to test whether protein and fat-depot signals share a causal variant; Steiger filtering used to assess direction of causal effect. |
| 8 | Sensitivity analyses and additional analyses | Describe any sensitivity or additional analyses performed. | 7-9 | MR-Egger, weighted median, simple mode, weighted mode; SIMEX correction for MR-Egger; MR-LASSO; MR-PRESSO; Radial MR; Cochran's Q; reverse two-sample MR; sex-stratified analyses; replication using MoBa discovery instruments; colocalisation; GTEx eQTL lookup; PheWAS. |
| 9a | Software and pre-registration | Name statistical software and package(s), including version and settings. | 9 | “All analyses were performed in R (version 4.2.0) using the ‘TwoSampleMR’, ‘RadialMR’, ‘MRPRESSO’, and ‘MendelianRandomization’ packages.” |
| 9b |  | State whether the study protocol was pre-registered (and when/where). | NA | This study was not pre-registered. |
| **RESULTS** | | | | |
| 10a | Descriptive data | Report numbers of individuals at each stage and reasons for exclusion; consider a flow diagram. | 11-18 | Candidate protein flow reported: 2,940 proteins tested in Step 1 → 1,995 excluded (no evidence of cBMI effect) → 945 remaining → 805 further excluded (temporally inconsistent or consistently significant across all timepoints) → 140 candidates entered Step 2 → 7 final directionally consistent mediators (3 for ASAT, 5 for GFAT, 1 overlapping). Figure 1 is referenced as an overview of the analytic design. |
| 10b |  | Report summary statistics for exposure(s), outcome(s) and other relevant variables. | 11 | Table 1 (referenced): ALSPAC cBMI sample N=736–5,780 across 12 timepoints, ~50/50 sex split (up to 55.4% female by age 18), median BMI rising from ~16.4–16.5 kg/m² (age 3) to ~21.8–22.1 kg/m² (age 18); UK Biobank fat-depot outcome sample predominantly White British, mean age ~64–65 years, mean BMI 26.0 kg/m² (female) / 27.1 kg/m² (male); UKB-PPP proteomic sample N=54,219, mean age 56.7 years, 54.3% female, 94.4% White. |
| 10c |  | If data sources include meta-analyses of previous studies, provide assessments of heterogeneity across studies. | NA | Not applicable — this study does not meta-analyse summary estimates across multiple previous studies; all summary statistics are drawn from single source GWAS per trait. |
| 10d |  | For two-sample MR: (i) justify similarity of variant–exposure/variant–outcome associations between samples; (ii) report sample overlap. | 24-25 | Discussed as a limitation: possible sample overlap between UKB-PPP (pQTL GWAS) and UK Biobank imaging-derived ASAT/GFAT GWAS, as both draw on overlapping UK Biobank participants; degree of overlap “cannot be precisely quantified”. By contrast, cBMI instrument-derivation cohorts (MoBa/ALSPAC) and the outcome cohort (UK Biobank) are stated to be non-overlapping. |
| 11a | Main results | Report associations between genetic variant and exposure, and between variant and outcome, on an interpretable scale. | 5,6,11 | Full SNP-level exposure and outcome association statistics are provided in Supplementary Tables 1 (cBMI instruments) and 2 (pQTL instruments); not reproduced in full in the main text. |
| 11b |  | Report MR estimates of the exposure–outcome relationship with measures of uncertainty, on an interpretable scale. | 11-18 | Reported throughout Results with beta coefficients and 95% CIs, e.g. cBMI on ASAT at age 7: beta=0.13 (95% CI 0.05, 0.21); cBMI on GFAT at age 8: beta=0.42 (95% CI 0.27, 0.56); protein–depot associations in Table 3 (e.g. ACAN–ASAT beta=−0.15, 95% CI −0.27, −0.04). |
| 11c |  | If relevant, translate relative risk estimates into absolute risk for a meaningful time period. | NA | Not applicable — outcomes are continuous MRI-derived fat-depot volumes rather than binary disease endpoints, so translation to absolute risk does not apply. |
| 11d |  | Consider plots to visualise results (forest plot, scatterplot of variant–outcome vs variant–exposure, etc.). | 11-18 | Figure 2 (primary MR associations across timepoints); Figure 3 (hierarchical clustering of candidate protein effects); Figure 4 (volcano plots of protein effects on ASAT and GFAT); Figure 5 (final directionally consistent candidate mediators). |
| 12a | Assessment of assumptions | Report the assessment of the validity of the assumptions. | 11-18 | F-statistics reported for instrument strength (median 8.13, range 4.91–10.78 across cBMI timepoints); colocalisation posterior probabilities reported for each candidate protein–depot pair; GTEx eQTL and PheWAS used as supporting mechanistic evidence; Steiger filtering performed as a directionality check. |
| 12b |  | Report additional statistics (e.g. heterogeneity assessments such as I², Q statistic, E-value). | 11-18 | Cochran's Q statistic used to quantify heterogeneity across instruments (referenced in Statistical and sensitivity analyses). Colocalisation posterior probabilities (H0–H4) reported: H1 range 0.92–0.93, H4 range 0.018–0.019 across all eight protein–fat-depot pairs. |
| 13a | Sensitivity analyses and additional analyses | Report sensitivity analyses assessing robustness to violations of assumptions. | 11-18 | Detailed by age band: weak/null and inconsistent associations across methods at ages 3–6 years; IVW consistently positive from age 7–14 years with concordant weighted-median/mode estimates and directionally consistent SIMEX-corrected MR-Egger; less consistent (but directionally similar) at ages 15–18, particularly for GFAT. Radial MR, MR-LASSO and MR-PRESSO estimates broadly concordant with IVW; MR-PRESSO-flagged outliers did not materially alter direction of effect. |
| 13b |  | Report results from other sensitivity or additional analyses. | 13-14 | Sex-stratified IVW analyses broadly consistent in direction with combined-sex results, with stronger/more precise estimates in females for both depots (Supplementary Table 8). Replication using stronger MoBa discovery instruments (F-stat ~33–38) reproduced timing and direction of primary ASAT/GFAT findings (Supplementary Table 9); VAT/liver/pancreas fat estimates remained null. |
| 13c |  | Report any assessment of direction of causal relationship (e.g. bidirectional MR). | 17-18 | Reverse two-sample MR reported: higher ASAT associated with lower circulating ACAN and higher CCL7 (suggesting possible bidirectionality); no reverse effect for CLIC5 on ASAT; borderline reciprocal association of CLIC5 with GFAT; reverse association between GFAT and CLEC10A; no reverse effects for BMP10, KIT or IGSF3. Steiger filtering performed as an additional directionality check (no exposure SNPs more strongly associated with the outcome). |
| 13d |  | When relevant, report and compare with estimates from non-MR analyses. |  | Not directly compared with non-MR (observational) estimates within a formal statistical framework in this study; comparisons with prior observational and genetic literature are discussed narratively in the Discussion (“Comparison with other studies”). |
| 13e |  | Consider additional plots (e.g. leave-one-out analyses). | NA | Considered, but not included in this instance. |
| **DISCUSSION** | | | | |
| 14 | Key results | Summarise key results with reference to study objectives. | 20 | Discussion opens by summarising that higher genetically predicted cBMI from age 7 years onward was causally associated with increased adult ASAT and GFAT but not VAT, liver, or pancreatic fat, and that two-step MR identified seven directionally and temporally consistent candidate mediating proteins (three for ASAT, five for GFAT, one overlapping). |
| 15 | Limitations | Discuss limitations, including validity of IV assumptions, other sources of bias, and imprecision; discuss direction/magnitude of potential bias. | 23-25 | Covered under “Strengths and limitations of this study”: variable/weak instrument strength across cBMI timepoints; inability to disentangle direct cBMI effects from those mediated via adult adiposity (MVMR not feasible); assumption of stable genetic architecture of BMI when applying MoBa-derived instruments across a wider ALSPAC age range; limitations of circulating vs tissue-specific protein measures; possible UKB-PPP/imaging-GWAS sample overlap; restriction to European-ancestry participants. |
| 16a | Interpretation | Meaning: give a cautious overall interpretation in context of limitations and comparison with other studies. | 20-23 | Discussion frames findings as depot-specific rather than generalised, with caveats threaded throughout (weak early-life instruments, MVMR not performed, possible sample overlap, bidirectionality for some proteins). |
| 16b |  | Mechanism: discuss underlying biological mechanisms and whether the gene–environment equivalence assumption is reasonable; use causal language carefully. | 18-23 | Discussed extensively under “Comparison with other studies” and “Exploration of candidate proteins”: ACAN (extracellular matrix proteoglycan) proposed to affect ASAT via ECM scaffolding; CCL7 (monocyte/macrophage recruitment) and CLIC5 (cytoskeletal/membrane integrity) proposed as immune/cytoskeletal mediators; BMP10 (vascular/developmental signalling), CLEC10A (antigen-presenting cell receptor), KIT (haematopoietic receptor tyrosine kinase) and IGSF3 (cell adhesion) proposed as GFAT mediators, supported by GTEx eQTL and PheWAS evidence. |
| 16c |  | Clinical relevance: discuss clinical/public policy relevance and implications for intervention effect sizes. | 25-26 | Covered under “Implications of findings”: mid-childhood (~age 7) proposed as a critical prevention window; contrasted with recent approval of GLP-1 receptor agonists for adolescents ≥12 years, suggesting earlier preventive intervention may be more impactful than later treatment; candidate proteins proposed as potential biomarkers/targets for functional follow-up studies. |
| 17 | Generalisability | Discuss generalisability (a) to other populations, (b) across other exposure periods/timings, (c) across other levels of exposure. | 23-25 | (a) Restricted to predominantly European-ancestry participants, explicitly flagged as limiting generalisability given known differences in BMI distribution, genetic architecture and cardiometabolic risk across ancestries. (b) Addressed via the assumption of partial stability in cBMI genetic architecture when MoBa-derived (birth–8 years) instruments are applied across the wider ALSPAC age range (3–18 years). |
| **OTHER INFORMATION** | | | | |
| 18 | Funding | Describe sources of funding and the funders' role, including funding for the original data sources. | 27-28 | Provided under “declarations” section. |
| 19 | Data and data sharing | Provide/reference where data can be accessed; state whether statistical code is publicly available and where. | 28 | Provided under “declarations” section. |
| 20 | Conflicts of interest | All authors should declare all potential conflicts of interest. | 28 | Provided under “declarations” section. |
